# Concern seriousness quantified in regulatory decisions as a main factor in questioning safe, trustworthy medical practice: a cross-sectional study

**DOI:** 10.64898/2026.11.09.26366477

**Authors:** Javier A. Caballero

## Abstract

**Background:** The main factors related to regulatory decision outcomes reflect the pragmatic expectations of physicians to deliver safe patient care and uphold trust in the profession. Yet empirically understanding critical ones, like concern seriousness, has proven difficult.

**Methods:** In this cross-sectional study, we developed a new quantitative measure of concern seriousness via expert focus groups. Using multivariate modelling, we investigated its statistical association with regulatory decisions about all physicians involved in the UK over the 10 years ending with March 2023, within a population of 416,610. Decisions by stage were: whether to investigate complaints; investigation outcomes, including adjudication hearing referral; and hearing outcomes. We controlled for dozens of potential confounders, simultaneously addressing many live questions; giving adjusted odds ratios for associations of outcomes to: criminal conviction, seriousness, allegation types, complaint sources, practice type (*e*.*g*. GP, paediatrician, trainee), workplace history, personal characteristics (*e*.*g*. age, sex), international/domestic qualification, socioeconomics (*e*.*g*. childhood deprivation), exam results (*e*.*g*. IELTS), and many more.

**Results:** Severe outcomes predominantly related to factors similar across stages: criminal conviction, high seriousness, allegations of criminality, honesty or fairness, and the complaint coming from the physician’s employer, police, or others acting in a professional capacity. Controlling for seriousness chiefly adjusted associations to conviction and allegation types, explaining some away.

**Conclusions:** All else equal, the seriousness of concerns was one of the main factors related to regulatory outcomes. Such main factors underline what is practically expected of physicians to avoid for safe and trustworthy medical practice.

## Introduction

Understanding the factors in severe disciplinary, legal, and regulatory decision outcomes for physicians is critical to clarifying what is pragmatically expected of them to keep practice safe and maintain public trust. Among the many interconnected variables associated with outcomes, ^1–19^ seriousness is qualitatively considered central, ^20^ but demonstrating this quantitatively remains challenging. Further, concerns are usually assumed serious if they led to severe outcomes, ^4,10,18,21^ and such circular reasoning obstructs answering whether serious concerns beget severe outcomes. ^22^ To address this, we created a measure of seriousness of concerns related to medical practice, then studied it alongside dozens of other variables potentially related to regulatory decisions about physicians in the UK.

Our approach draws parallels with patient case severity-scoring systems routinely used in many medical areas. ^23,24^ It also conceptually aligns with crime severity, seriousness, or harm measures implemented worldwide ^25–33^ to help focus resources on the cases of highest value for public protection. ^33^ We extended these to ranking non-criminal matters alongside criminal ones, encompassing all that is historically coded in the UK as potentially affecting physicians’ practice.

We found that severe regulatory decision outcomes about physicians followed serious concerns. By high-lighting which factors were or were not related to such decisions, our findings can help prioritise limited resources to enhance policy, governance, education, and workplace cultures across similar systems globally.

## Methods

Anyone can file a complaint about a physician with the General Medical Council (GMC), which may go through up to three decision stages, as follows. A “triage” decision on whether to investigate. If so, an investigation decision with potential outcomes: no further action, letter of advice, warning, undertaking, or referral to an adjudication hearing at the Medical Practitioners Tribunal Service. If the latter, hearing decision outcomes may be: no further action (no sanction or warning), warning, undertaking or conditions (limitation of practice), suspension (of the right to practise), or erasure (its loss). Correspondingly, the most severe outcomes are: investigation, referral to a hearing, and erasure.

### Data sourcing and preparation

We engineered three datasets from the GMC’s databases —one per decision stage. For comparability, independence, and to guard against bias, we used data on all decisions made in the 10 years up to 31-March-2023, about the first ever complaint, investigation, or hearing about identified physicians (tables 1 and 2).

**Table 1:**
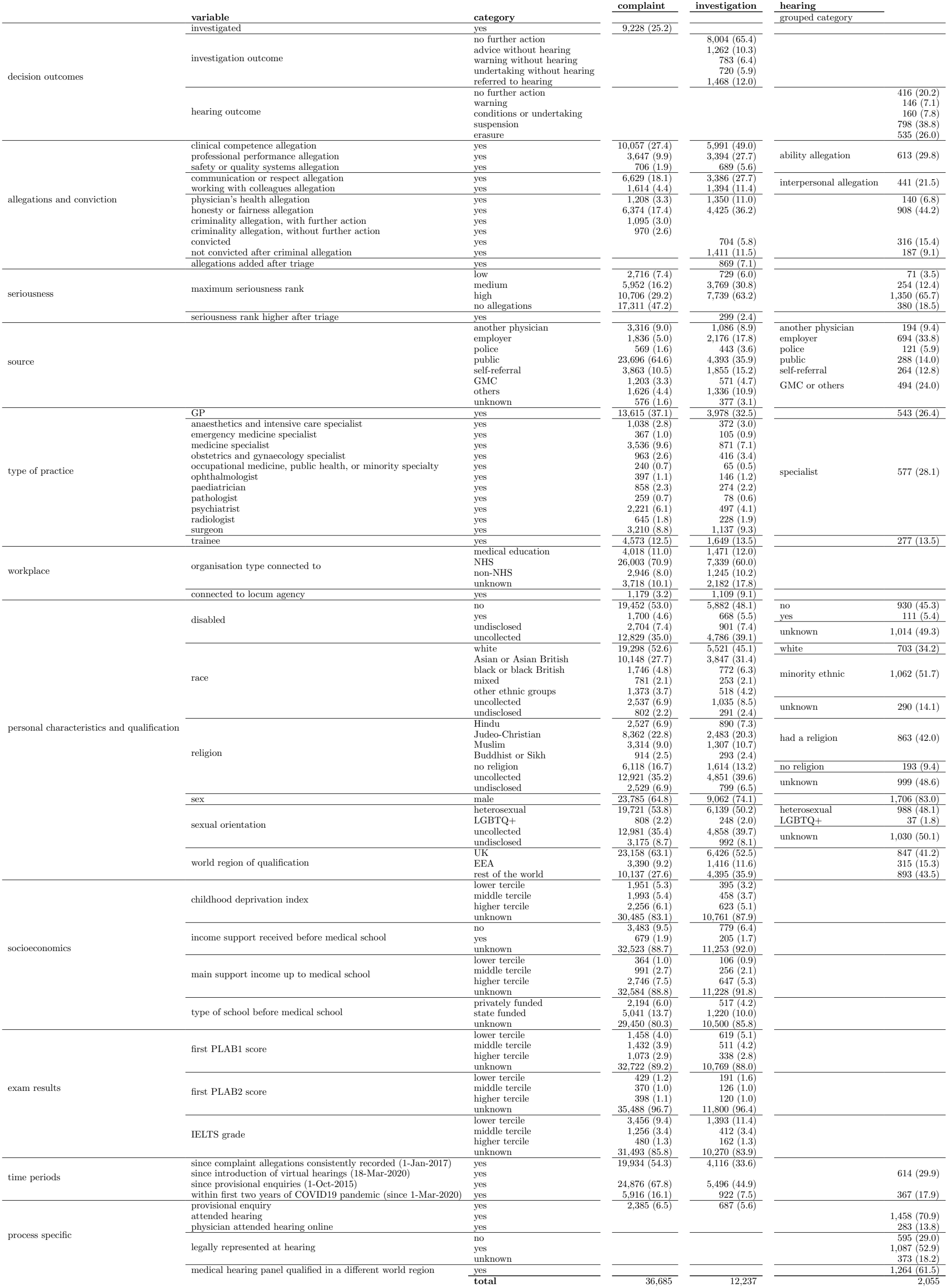
Summary of categorical variables: count (% of total). Only one category is shown for binary variables. Variables consolidated further for first hearings are named in the “grouped category” column.

|  |  |  | **complaint** | **investigation** | **hearing** |  |
|  | **variable** | **category** |  |  | grouped category |  |
|---|---|---|---|---|---|---|
| decision outcomes | investigated | yes | 9,228 (25.2) |  |  |  |
|  | investigation outcome | no further action |  | 8,004 (65.4) |  |  |
|  |  | advice without hearing |  | 1,262 (10.3) |  |  |
|  |  | warning without hearing |  | 783 (6.4) |  |  |
|  |  | undertaking without hearing |  | 720 (5.9) |  |  |
|  |  | referred to hearing |  | 1,468 (12.0) |  |  |
|  | hearing outcome | no further action |  |  |  | 416 (20.2) |
|  |  | warning |  |  |  | 146 (7.1) |
|  |  | conditions or undertaking |  |  |  | 160 (7.8) |
|  |  | suspension |  |  |  | 798 (38.8) |
|  |  | erasure |  |  |  | 535 (26.0) |
| allegations and conviction | clinical competence allegation | yes | 10,057 (27.4) | 5,991 (49.0) | ability allegation | 613 (29.8) |
|  | professional performance allegation | yes | 3,647 (9.9) | 3,394 (27.7) |  |  |
|  | safety or quality systems allegation | yes | 706 (1.9) | 689 (5.6) |  |  |
|  | communication or respect allegation | yes | 6,629 (18.1) | 3,386 (27.7) | interpersonal allegation | 441 (21.5) |
|  | working with colleagues allegation | yes | 1,614 (4.4) | 1,394 (11.4) |  |  |
|  | physician's health allegation | yes | 1,208 (3.3) | 1,350 (11.0) |  | 140 (6.8) |
|  | honesty or fairness allegation | yes | 6,374 (17.4) | 4,425 (36.2) |  | 908 (44.2) |
|  | criminality allegation, with further action | yes | 1,095 (3.0) |  |  |  |
|  | criminality allegation, without further action | yes | 970 (2.6) |  |  |  |
|  | convicted | yes |  | 704 (5.8) |  | 316 (15.4) |
|  | not convicted after criminal allegation | yes |  | 1,411 (11.5) |  | 187 (9.1) |
|  | allegations added after triage | yes |  | 869 (7.1) |  |  |
| seriousness | maximum seriousness rank | low | 2,716 (7.4) | 729 (6.0) |  | 71 (3.5) |
|  |  | medium | 5,952 (16.2) | 3,769 (30.8) |  | 254 (12.4) |
|  |  | high | 10,706 (29.2) | 7,739 (63.2) |  | 1,350 (65.7) |
|  |  | no allegations | 17,311 (47.2) |  |  | 380 (18.5) |
|  | seriousness rank higher after triage | yes |  | 299 (2.4) |  |  |
| source |  | another physician | 3,316 (9.0) | 1,086 (8.9) | another physician | 194 (9.4) |
|  |  | employer | 1,836 (5.0) | 2,176 (17.8) | employer | 694 (33.8) |
|  |  | police | 569 (1.6) | 443 (3.6) | police | 121 (5.9) |
|  |  | public | 23,696 (64.6) | 4,393 (35.9) | public | 288 (14.0) |
|  |  | self-referral | 3,863 (10.5) | 1,855 (15.2) | self-referral | 264 (12.8) |
|  |  | GMC | 1,203 (3.3) | 571 (4.7) | GMC or others | 494 (24.0) |
|  |  | others | 1,626 (4.4) | 1,336 (10.9) |  |  |
|  |  | unknown | 576 (1.6) | 377 (3.1) |  |  |
| type of practice | GP | yes | 13,615 (37.1) | 3,978 (32.5) |  | 543 (26.4) |
|  | anaesthetics and intensive care specialist | yes | 1,038 (2.8) | 372 (3.0) | specialist | 577 (28.1) |
|  | emergency medicine specialist | yes | 367 (1.0) | 105 (0.9) |  |  |
|  | medicine specialist | yes | 3,536 (9.6) | 871 (7.1) |  |  |
|  | obstetrics and gynaecology specialist | yes | 963 (2.6) | 416 (3.4) |  |  |
|  | occupational medicine, public health, or minority specialty | yes | 240 (0.7) | 65 (0.5) |  |  |
|  | ophthalmologist | yes | 397 (1.1) | 146 (1.2) |  |  |
|  | paediatrician | yes | 858 (2.3) | 274 (2.2) |  |  |
|  | pathologist | yes | 259 (0.7) | 78 (0.6) |  |  |
|  | psychiatrist | yes | 2,221 (6.1) | 497 (4.1) |  |  |
|  | radiologist | yes | 645 (1.8) | 228 (1.9) |  |  |
|  | surgeon | yes | 3,210 (8.8) | 1,137 (9.3) |  |  |
|  | trainee | yes | 4,573 (12.5) | 1,649 (13.5) |  | 277 (13.5) |
| workplace | organisation type connected to | medical education | 4,018 (11.0) | 1,471 (12.0) |  |  |
|  |  | NHS | 26,003 (70.9) | 7,339 (60.0) |  |  |
|  |  | non-NHS | 2,946 (8.0) | 1,245 (10.2) |  |  |
|  |  | unknown | 3,718 (10.1) | 2,182 (17.8) |  |  |
|  | connected to locum agency | yes | 1,179 (3.2) | 1,109 (9.1) |  |  |
| personal characteristics and qualification | disabled | no | 19,452 (53.0) | 5,882 (48.1) | no | 930 (45.3) |
|  |  | yes | 1,700 (4.6) | 668 (5.5) | yes | 111 (5.4) |
|  |  | undisclosed | 2,704 (7.4) | 901 (7.4) | unknown | 1,014 (49.3) |
|  |  | uncollected | 12,829 (35.0) | 4,786 (39.1) |  |  |
|  | race | white | 19,298 (52.6) | 5,521 (45.1) | white | 703 (34.2) |
|  |  | Asian or Asian British | 10,148 (27.7) | 3,847 (31.4) | minority ethnic | 1,062 (51.7) |
|  |  | black or black British | 1,746 (4.8) | 772 (6.3) |  |  |
|  |  | mixed | 781 (2.1) | 253 (2.1) |  |  |
|  |  | other ethnic groups | 1,373 (3.7) | 518 (4.2) |  |  |
|  |  | uncollected | 2,537 (6.9) | 1,035 (8.5) | unknown | 290 (14.1) |
|  |  | undisclosed | 802 (2.2) | 291 (2.4) |  |  |
|  | religion | Hindu | 2,527 (6.9) | 890 (7.3) | had a religion | 863 (42.0) |
|  |  | Judeo-Christian | 8,362 (22.8) | 2,483 (20.3) |  |  |
|  |  | Muslim | 3,314 (9.0) | 1,307 (10.7) |  |  |
|  |  | Buddhist or Sikh | 914 (2.5) | 293 (2.4) |  |  |
|  |  | no religion | 6,118 (16.7) | 1,614 (13.2) | no religion | 193 (9.4) |
|  |  | uncollected | 12,921 (35.2) | 4,851 (39.6) | unknown | 999 (48.6) |
|  |  | undisclosed | 2,529 (6.9) | 799 (6.5) |  |  |
|  | sex | male | 23,785 (64.8) | 9,062 (74.1) |  | 1,706 (83.0) |
|  | sexual orientation | heterosexual | 19,721 (53.8) | 6,139 (50.2) | heterosexual | 988 (48.1) |
|  |  | LGBTQ+ | 808 (2.2) | 248 (2.0) | LGBTQ+ | 37 (1.8) |
|  |  | uncollected | 12,981 (35.4) | 4,858 (39.7) | unknown | 1,030 (50.1) |
|  |  | undisclosed | 3,175 (8.7) | 992 (8.1) |  |  |
|  | world region of qualification | UK | 23,158 (63.1) | 6,426 (52.5) |  | 847 (41.2) |
|  |  | EEA | 3,390 (9.2) | 1,416 (11.6) |  | 315 (15.3) |
|  |  | rest of the world | 10,137 (27.6) | 4,395 (35.9) |  | 893 (43.5) |
| socioeconomics | childhood deprivation index | lower tercile | 1,951 (5.3) | 395 (3.2) |  |  |
|  |  | middle tercile | 1,993 (5.4) | 458 (3.7) |  |  |
|  |  | higher tercile | 2,256 (6.1) | 623 (5.1) |  |  |
|  |  | unknown | 30,485 (83.1) | 10,761 (87.9) |  |  |
|  | income support received before medical school | no | 3,483 (9.5) | 779 (6.4) |  |  |
|  |  | yes | 679 (1.9) | 205 (1.7) |  |  |
|  |  | unknown | 32,523 (88.7) | 11,253 (92.0) |  |  |
|  | main support income up to medical school | lower tercile | 364 (1.0) | 106 (0.9) |  |  |
|  |  | middle tercile | 991 (2.7) | 256 (2.1) |  |  |
|  |  | higher tercile | 2,746 (7.5) | 647 (5.3) |  |  |
|  |  | unknown | 32,584 (88.8) | 11,228 (91.8) |  |  |
|  | type of school before medical school | privately funded | 2,194 (6.0) | 517 (4.2) |  |  |
|  |  | state funded | 5,041 (13.7) | 1,220 (10.0) |  |  |
|  |  | unknown | 29,450 (80.3) | 10,500 (85.8) |  |  |
| exam results | first PLAB1 score | lower tercile | 1,458 (4.0) | 619 (5.1) |  |  |
|  |  | middle tercile | 1,432 (3.9) | 511 (4.2) |  |  |
|  |  | higher tercile | 1,073 (2.9) | 338 (2.8) |  |  |
|  |  | unknown | 32,722 (89.2) | 10,769 (88.0) |  |  |
|  | first PLAB2 score | lower tercile | 429 (1.2) | 191 (1.6) |  |  |
|  |  | middle tercile | 370 (1.0) | 126 (1.0) |  |  |
|  |  | higher tercile | 398 (1.1) | 120 (1.0) |  |  |
|  |  | unknown | 35,488 (96.7) | 11,800 (96.4) |  |  |
|  | IELTS grade | lower tercile | 3,456 (9.4) | 1,393 (11.4) |  |  |
|  |  | middle tercile | 1,256 (3.4) | 412 (3.4) |  |  |
|  |  | higher tercile | 480 (1.3) | 162 (1.3) |  |  |
|  |  | unknown | 31,493 (85.8) | 10,270 (83.9) |  |  |
| time periods | since complaint allegations consistently recorded (1-Jan-2017) | yes | 19,934 (54.3) | 4,116 (33.6) |  |  |
|  | since introduction of virtual hearings (18-Mar-2020) | yes |  |  |  | 614 (29.9) |
|  | since provisional enquiries (1-Oct-2015) | yes | 24,876 (67.8) | 5,496 (44.9) |  |  |
|  | within first two years of COVID19 pandemic (since 1-Mar-2020) | yes | 5,916 (16.1) | 922 (7.5) |  | 367 (17.9) |
| process specific | provisional enquiry | yes | 2,385 (6.5) | 687 (5.6) |  |  |
|  | attended hearing | yes |  |  |  | 1,458 (70.9) |
|  | physician attended hearing online | yes |  |  |  | 283 (13.8) |
|  | legally represented at hearing | no |  |  |  | 595 (29.0) |
|  |  | yes |  |  |  | 1,087 (52.9) |
|  |  | unknown |  |  |  | 373 (18.2) |
|  | medical hearing panel qualified in a different world region | yes |  |  |  | 1,264 (61.5) |
|  |  | **total** | 36,685 | 12,237 |  | 2,055 |

**Table 2:** Summary of numerical variables: mean (standard deviation)

| **variable** | **mean (SD)** | **range** |
|---|---|---|
| *complaint* |  |  |
| age (years) | 46.6 (11.2) | 22.7 - 90.6 |
| allegations’ count | 1.3 (1.9) | 0.0 - 33.0 |
| count of organisations worked at | 2.5 (1.9) | 0.0 - 14.0 |
| fraction of lifetime with right to practise in the UK | 0.4 (0.2) | 0.0 - 0.7 |
| revalidation deferrals’ count | 0.2 (0.6) | 0.0 - 10.0 |
| *investigation* |  |  |
| age (years) | 47.6 (11.7) | 23.0 - 86.9 |
| allegations’ count | 3.3 (3.1) | 0.0 - 60.0 |
| count of organisations worked at | 2.1 (1.7) | 0.0 - 13.0 |
| fraction of lifetime with right to practise in the UK | 0.4 (0.2) | 0.0 - 0.7 |
| prior complaints’ count | 0.3 (1.0) | 0.0 - 83.0 |
| revalidation deferrals’ count | 0.2 (0.6) | 0.0 - 8.0 |
| *hearing* |  |  |
| age (years) | 47.3 (11.8) | 23.4 - 90.6 |
| allegations’ count | 2.6 (3.3) | 0.0 - 55.0 |
| fraction of panel of opposite sex | 0.5 (0.2) | 0.0 - 1.0 |

We grouped five ‘undertaking’ hearing outcomes with ‘conditions’ outcomes. We grouped 24 ‘voluntary erasure’ hearing outcomes (relinquishing registration) into ‘no further action’ outcomes.

Each allegation was logged using one of 684 codes. Each code belonged in one of nine mutually exclusive allegation types. Cases could have allegations of several types. For triage decisions, we subdivided criminality allegations by whether further action was allegedly taken. For investigation and hearing stages, we coded the availability of conviction documentation, separately coding criminality allegations without conviction. Allegations were just claims unless a hearing proved them with evidence. We only used data on proven allegations to study hearing decisions. The date of birth, sex, and place of qualification of physicians were verified upon registration. Other personal characteristics were self-reported. We used the Income Deprivation Affecting Children Index to measure childhood deprivation. The ‘main support income up to medical school’ was the tercile among the average salaries of all occupations in the Standard Occupational Classification (Office for National Statistics, UK), for the occupation of the highest-earning parent or guardian. Test scores were from the first GMC-recorded IELTS or Professional and Linguistic Assessments Board theoretical or practical test (PLAB1 or PLAB2).

All time-dependent variables were at the complaint date, including whether physicians were on the relevant GP or specialist registers, and the organisation type that physicians connected-to for revalidation. Birth date was missing for seven physicians referred to hearing; we imputed their age as the time between qualification and complaint, plus 24 years. The ‘fraction of lifetime with right to practise in the UK’ was the ratio of the time since first registering or becoming licensed (licensing became required in November 2009) and age. For 33 complaints and 12 investigations with missing data, we imputed the inverse of this fraction as the average of the 30 nearest neighbours of each observation, using all other variables controlled for in our models.

### Statistical methods

We analysed our data using bespoke Python (3.9.13) and R (4.3.1) scripts. Due to our large sample sizes, we reported those explicitly and used statistical testing sparingly. Unless unavailable, we favoured non-parametric methods.

We controlled for variables that addressed questions of interest, checked for collinearity, and *z*-scored numerical variables. Results came from three descriptive models of triage, investigation, and hearing decisions, plus comparing these to corresponding nested reduced models that lacked seriousness variables. For brevity, we omitted detailed results of these subsidiary reduced models.

We used logistic regression to model triage decisions (*c*-statistic 0.84, McFadden pseudo-*R*^2^ 0.28). We modelled investigation and hearing decisions with multinomial regressions, setting ‘no further action’ outcomes as the baseline (multi-class areas under the ROC curve 0.79 and 0.81, McFadden pseudo-*R*^2^ 0.24 and 0.28, respectively). For the smaller sample of hearing decisions, we considered only variables that were important at earlier stages or addressed questions of interest, grouping some into fewer categories. For a straightforward interpretation, we did not consider interactions between variables.

We set significance levels at *α* = 0.01 for the triage and investigation models, and *α* = 0.05 for hearing models, considering the smaller dataset. We fully reported modelling results yet focused on discussing the ~20 % of factors with the highest effect sizes, that were significant, since these provided the highest explanatory value. Automated bi-directional step-wise modelling, minimising the Akaike Information Criterion (AIC), confirmed the retention of such main factors.

### Measuring seriousness

Regulatory decision-makers gauge the seriousness of concerns in determining appropriate outcomes. ^20^ In five focus groups, we preliminarily ranked-ordered the 684 individual allegation codes by their seriousness, with two experts on regulatory decision data; followed by two subsequent sessions with them plus one expert in triage decisions and another in investigation decisions. Using a bubble-sort method, we iteratively traversed the list of codes, asking “Typically, how seriously would a physician’s fitness to practise be impaired if such an allegation was true?”, relocating codes until we reached consensus on the ordering of the whole list. Some codes were duplicates (older and newer terminology) or were arguably equally serious, like discrimination based on age or religion. This assigned an integer from 1 to 450 to each of the 684 codes (table 3). Achieving universal consensus is practically unattainable, since additional raters might rank single codes slightly differently. But if we agree that codes a few ranks apart are sensibly ordered, our ranking arguably remains valuable for quantitative analysis.

**Table 3:** Allegation ranking. Patient or colleague discrimination at ranks 323–324 could be about age, culture, disability, gender, lifestyle, pregnancy or maternity, race, religion or beliefs, sex, sexual orientation, socioeconomic status, or transgender status.

| allegation code | rank | allegation code | rank | allegation code | rank |
|---|---|---|---|---|---|
| murder | 450 | failure to examine | 300 | resisting arrest with no further action | 150 |
| murder conditional discharge | 449 | inadequate examination | 299 | lack of further investigation | 149 |
| murder caution | 448 | inadequate assessment | 298 | prompt treatment or investigation not undertaken | 148 |
| murder with no further action | 447 | poor communication skills | 297 | failure to arrange ongoing care | 147 |
| attempted murder | 446 | internet prescribing | 296 | delegation to inappropriate physician or other | 146 |
| attempted murder caution | 445 | withholding treatment | 295 | rough handling of patient | 145 |
| attempted murder with no further action | 444 | care prejudiced by complaint | 294 | no handover to clinician or team | 144 |
| manslaughter | 443 | communication in care prejudiced by complaint | 293 | failure to arrange adequate cover | 143 |
| manslaughter conditional discharge | 442 | withholding referral | 292 | failure to rectify harm | 142 |
| manslaughter caution | 441 | failure to refer when appropriate | 291 | failure to prioritise clinical need | 141 |
| manslaughter with no further action | 440 | inappropriate referral | 290 | failure to respect patient’s dignity | 140 |
| rape | 439 | inappropriate expression of beliefs | 289 | failure to respect patient’s views | 139 |
| rape conditional discharge | 438 | inappropriate refusal to provide care | 288 | failure to visit | 138 |
| rape caution | 437 | inappropriate delay in providing care | 287 | no alternative offered after conscientious objection | 137 |
| rape with no further action | 436 | inadequate knowledge of the English language | 286 | soliciting | 136 |
| female circumcision | 435 | failure to meet language needs | 285 | soliciting conditional discharge | 135 |
| female circumcision caution | 434 | unnecessary risk to research participants | 284 | soliciting caution | 134 |
| female circumcision with no further action | 433 | irresponsible supplying of medicine | 283 | soliciting with no further action | 133 |
| sexual assault | 432 | failure to offer appropriate help in emergencies | 282 | failure to work in partnership | 132 |
| sexual assault with no further action | 431 | not accessible when on duty | 281 | failure to share information with colleague | 131 |
| indecent assault | 430 | failure to take up post | 280 | failure to keep colleagues informed about care continuity | 130 |
| indecent assault conditional discharge | 429 | failure to comply with Performance Assessment | 279 | failure to keep colleagues informed about care | 129 |
| indecent assault caution | 428 | failure to comply with Health Assessment | 278 | failure to share information appropriately | 128 |
| attempted rape | 427 | failure to comply with Language Assessment | 277 | failure to ensure basic care | 127 |
| attempted rape with no further action | 426 | failure to cooperate with investigation | 276 | no consent for research | 126 |
| making indecent images of children | 425 | failure to cooperate with inquest or inquiry | 275 | other failure to follow regulator’s guidance | 125 |
| making indecent images of children caution | 424 | failure to tell regulator about criminal conviction | 274 | failure to respond to risk with possible abuse | 124 |
| making indecent images of children with no further action | 423 | leaving post early or without notice | 273 | public disorder | 123 |
| child pornography offences | 422 | no indemnity insurance | 272 | public disorder with no further action | 122 |
| child pornography offences conditional discharge | 421 | failure to tell regulator about criminal charge | 271 | failure to raise concern about patient safety | 121 |
| child pornography offences caution | 420 | failure to tell regulator about criminal offence | 270 | failure to maintain confidentiality of patient | 120 |
| child pornography offences with no further action | 419 | delay in telling regulator about criminal charge or offence | 269 | insufficient information on referral | 119 |
| possessing indecent images of children caution | 418 | driving under the influence of alcohol | 268 | ignoring colleague’s advice | 118 |
| improper relationship with patient | 417 | driving under the influence of alcohol conditional discharge | 267 | delay in reporting colleague risk | 117 |
| indecent exposure | 416 | driving under the influence of alcohol caution | 266 | failure to get second opinion | 116 |
| indecent exposure with no further action | 415 | driving under the influence of alcohol with no further action | 265 | refuse patient second opinion | 115 |
| Indecent behaviour | 414 | driving under the influence of drugs | 264 | insufficient information on delegation | 114 |
| breach of non-molestation order | 413 | driving under the influence of with no further action | 263 | breach of information security | 113 |
| breach of non-molestation order with no further action | 412 | failure to provide a specimen | 262 | failure to provide adequate systems to manage risk | 112 |
| voyeurism | 411 | failure to provide a specimen with no further action | 261 | failure to report inadequate systems to manage risk | 111 |
| voyeurism with no further action | 410 | intentional breach of information security | 260 | inadequate practice arrangements | 110 |
| inappropriate examination | 409 | intentional disclosure of confidential information | 259 | failure to respond to complaints | 109 |
| work when suspended by regulator | 408 | failure to seek health investigation or treatment | 258 | inappropriately providing care to family or friends | 108 |
| failure to tell patient about practice restriction | 407 | suitable action not taken | 257 | failure to meet communication needs | 107 |
| dishonesty with patient or colleague | 406 | failure to remedy performance deficiencies | 256 | refuse to get second opinion | 106 |
| dishonesty with patient | 405 | failure to remedy teamwork deficiencies | 255 | false or misleading advertising | 105 |
| terrorism offences | 404 | failure to clarify own limits | 254 | undeclared conflict of interest in home clinic | 104 |
| terrorism offences caution | 403 | failure to change practice | 253 | other undeclared conflict of interest | 103 |
| terrorism offences with no further action | 402 | failure to recognise own limits | 252 | undeclared conflict of interest in hospital | 102 |
| assisting suicide | 401 | failure to follow regulatory guidance | 251 | failure to make a reasonable adjustment | 101 |
| assisting suicide with no further action | 400 | failure to diagnose | 250 | failure to arrange alternative to access care | 100 |
| offences under Abortion Act | 399 | self treating or prescribing | 249 | failure to verify before signing report | 99 |
| offences under Abortion Act caution | 398 | treatment or prescription for family or friends | 248 | inappropriate personal comments | 98 |
| offences under Abortion Act with no further action | 397 | treatment or prescription for family | 247 | inappropriate factors taken into account for care decision | 97 |
| assault with grievous bodily harm | 396 | breach of regulations regarding home clinic | 246 | health and safety breach | 96 |
| assault with grievous bodily harm conditional discharge | 395 | breach of home clinic regulations with no further action | 245 | failure to participate in audit | 95 |
| assault with grievous bodily harm caution | 394 | conduct on breach of regulations regarding home clinic | 244 | failure to report risk | 94 |
| assault with grievous bodily harm with no further action | 393 | performance related to breach of regulations regarding home clinic | 243 | not consulting colleagues | 93 |
| false certifications | 392 | failure to provide name or registration number | 242 | failure to appropriately supervise in teaching, training, or assessment | 92 |
| improper alteration of records | 391 | failure to seek independent advice | 241 | failure to appropriately supervise | 91 |
| false claims to qualifications | 390 | unfairly ending relationship with prejudice | 240 | financial maladministration of residential home | 90 |
| false claims to regulatory status | 389 | unfairly ending relationship | 239 | financial maladministration of nursing home | 89 |
| false claims to experience | 388 | failure to protect patients from Hepatitis B | 238 | motoring offences | 88 |
| assault causing actual bodily harm | 387 | tuberculosis | 237 | motoring offences with no further action | 87 |
| assault causing actual bodily harm conditional discharge | 386 | ICD F16: abuse of hallucinogens | 236 | minor motoring offences | 86 |
| assault causing actual bodily harm caution | 385 | ICD F20-29: schizophrenia, etc. | 235 | minor motoring offences conditional discharge | 85 |
| assault causing actual bodily harm with no further action | 384 | ICD F19: multiple substance abuse | 234 | minor motoring offences caution | 84 |
| theft | 383 | ICD F90-98: behavioural disorders | 233 | other offences | 83 |
| theft conditional discharge | 382 | ICD F50-59: other behavioural disorders, etc. | 232 | other offences with no further action | 82 |
| theft caution | 381 | ICD F11: abuse of opioids | 231 | failure to assist regarding possible abuse | 81 |
| theft with no further action | 380 | ICD F14: abuse of cocaine | 230 | failure to arrange ongoing care after relationship ends | 80 |
| arson | 379 | ICD F15: abuse of stimulants | 229 | failure to respect advance wishes | 79 |
| firearms offences | 378 | ICD F13: abuse of sedatives or hypnotics | 228 | verbal abuse towards patient | 78 |
| firearms offences caution | 377 | ICD F00-09: organic mental disorders, etc. | 227 | failure to share appropriate information with patient’s family or friends | 77 |
| common assault | 376 | ICD F30-39: affective disorders | 226 | failure to optimise pain relief | 76 |
| common assault conditional discharge | 375 | ICD F60-69: personality disorders, etc. | 225 | poor relations with colleagues | 75 |
| common assault caution | 374 | ICD F80-89: psychological development | 224 | failure to cooperate with complaint procedure | 74 |
| common assault with no further action | 373 | ICD F40-49: neuroses, etc. | 223 | failure to ask for appropriate advice or information | 73 |
| affray | 372 | ICD F10: alcohol abuse | 222 | failure to offer apology | 72 |
| affray with no further action | 371 | ICD F18: abuse of solvents | 221 | failure to explain error or issue | 71 |
| perjury | 370 | ICD F12: abuse of cannabinoids | 220 | failure to reflect on practice to maintain performance | 70 |
| perjury caution | 369 | ICD F17: abuse of tobacco | 219 | failure to reflect on practice to protect patients | 69 |
| perjury with no further action | 368 | other mental or behavioural disorder | 218 | failure to respond to communication concerns | 68 |
| perverting course of justice | 367 | other physical disorder | 217 | failure to offer appropriate help in responding to safety risks | 67 |
| perverting course of justice with no further action | 366 | failure to seek independent health advice | 216 | no assessment of patient’s priorities | 66 |
| forgery | 365 | failure to seek independent medical care | 215 | failure to provide relevant information systems to protect patients | 65 |
| practising without licence | 364 | chronic fatigue syndrome | 214 | failure to work in partnership with patient | 64 |
| fraud | 363 | ICD H53.9: unspecified visual disturbance | 213 | failure to respect patient’s views | 63 |
| bullying colleague physically and/or verbally | 362 | ICD H90.2: unspecified loss of conductive hearing | 212 | encouraging gifts or donations | 62 |
| bullying colleague physically | 361 | inadequate knowledge of the law or legal codes | 211 | pressurising patients | 61 |
| bullying colleague verbally | 360 | inappropriate or irresponsible prescribing | 210 | failure to maintain trust socially or medically | 60 |
| fraud conditional discharge | 359 | prescribing without adequate history | 209 | inappropriate use of information in research | 59 |
| fraud caution | 358 | chaperone not present during examination | 208 | inaccurate reference or appraisal on teaching, training, or assessing | 58 |
| fraud with no further action | 357 | impaired by barring decision in England | 207 | inaccurate reference or report | 57 |
| false or misleading evidence | 356 | failure to protect patients from other communicable diseases | 206 | failure to listen to patient | 56 |
| dishonest reference, appraisal, or report | 355 | failure to protect colleagues from disease | 205 | insufficient information for consent to treatment | 55 |
| false or misleading research report | 354 | failure to protect patients from other diseases | 204 | insufficient information for consent to research | 54 |
| false or misleading report | 353 | misuse of funds | 203 | insufficient information for consent to screening | 53 |
| false or misleading reference | 352 | inappropriate use of medical records in research | 202 | failure to support colleagues during teaching, training, or assessing | 52 |
| given misleading evidence | 351 | failure to report adverse reaction | 201 | not keeping patient’s GP informed | 51 |
| dishonesty in assessment or appraisal | 350 | inadequate follow up | 200 | unclear about responsibilities | 50 |
| work when suspended from performers’ list | 349 | prescribing without examination | 199 | no information or evidence folder | 49 |
| research fraud | 348 | possessing an offensive weapon | 198 | failure to participate in team review | 48 |
| drug trial fraud | 347 | possessing an offensive weapon conditional discharge | 197 | failure to inform participant’s physician about research | 47 |
| ethical approval not obtained for research | 346 | possessing an offensive weapon caution | 196 | insufficient information given about examination | 46 |
| use of inappropriate participants in research | 345 | possessing an offensive weapon with no further action | 195 | failure to justify decision to end relationship | 45 |
| financial deception | 344 | coercion of possible participants in research | 194 | failure to provide appropriate information to patient | 44 |
| offering inducements | 343 | minor dishonesty offence | 193 | delay in handover of records | 43 |
| accepting inducements | 342 | minor dishonesty offence caution | 192 | accidental breach of information security | 42 |
| interest conflict affecting patient care | 341 | breach of confidentiality in social media | 191 | rudeness to patient | 41 |
| failure to declare interest | 340 | failure to tell regulator about caution | 190 | causing employment issues to colleagues | 40 |
| false claims to membership | 339 | coercive controlling behaviour | 189 | unclear about responsibilities in a team | 39 |
| failure to tell employer of regulatory restriction | 338 | coercive controlling behaviour with no further action | 188 | inappropriate IT equipment use | 38 |
| threats to kill | 337 | failure to get necessary performance training | 187 | poor assessment or appraisal during teaching, training, or assessing | 37 |
| threats to kill conditional discharge | 336 | failure to get necessary maintenance or improvement training | 186 | poor appraisal | 36 |
| threats to kill caution | 335 | failure to report unsafely practising colleague | 185 | poor assessment | 35 |
| threats to kill with no further action | 334 | inaccurate medical records | 184 | poor teaching, training, or assessing | 34 |
| harassment with obscene material | 333 | cruelty to animals | 183 | poor teaching or supervision skills | 33 |
| racial harassment | 332 | causing unnecessary suffering to a dog | 182 | accidental disclosure of confidential information | 32 |
| harassment | 331 | failure to declare financial or commercial interest | 181 | failure to be objective in teaching, training, assessment, or appraisal | 31 |
| harassment caution | 330 | failure to pay fees to another physician | 180 | illegible medical records | 30 |
| harassment with no further action | 329 | inadequate participation in medical education | 179 | failure to appropriate audit or review on patient safety systems | 29 |
| controlled substances offences | 328 | inadequate participation in medical education to keep up-to-date | 178 | failure to appropriate audit or review on teaching or supervision | 28 |
| controlled substances offences conditional discharge | 327 | unjustifiable service claims | 177 | failure to respect others’ skills | 27 |
| controlled substances offences caution | 326 | drunk and disorderly | 176 | failure to tell fees to another physician | 26 |
| controlled substances offences with no further action | 325 | drunk and disorderly conditional discharge | 175 | poor reaction to adverse event | 25 |
| patient discrimination | 324 | drunk and disorderly caution | 174 | poor communication with young or old patients | 24 |
| colleague discrimination | 323 | inadequate leadership | 173 | failure to participate in quality assurance | 23 |
| wilfully abandoning child younger than 16 | 322 | failure to provide information about fees | 172 | delay in providing report or documentation | 22 |
| wilfully abandoning child younger than 16 conditional discharge | 321 | inappropriate use of information in research | 171 | failure to explain to patients information sharing | 21 |
| wilfully abandoning child younger than 16 with no further action | 320 | diabetes mellitus with complications | 170 | inappropriate canvassing | 20 |
| harassing colleagues | 319 | delay in diagnosis | 169 | failure to support colleagues | 19 |
| not declaring regulatory actions at licence application | 318 | maladministration of nursing or residential home | 168 | lack of commitment | 18 |
| delay telling employer of practice restriction | 317 | incomplete medical records | 167 | failure to understand educational principles | 17 |
| delay telling patient of practice restriction | 316 | serious motoring offences | 166 | unresponsive to individual needs in teaching or supervision | 16 |
| inadequate knowledge base | 315 | serious motoring offences conditional discharge | 165 | failure to promote children’s health | 15 |
| misdiagnosis | 314 | serious motoring offences caution | 164 | failure to promote patient health | 14 |
| consent issues on patient lacking capacity | 313 | driving without tax, insurance, or licence | 163 | failure to properly inform patient about end of relationship | 13 |
| fairness issues about patient lacking capacity | 312 | driving without tax, insurance, or licence conditional discharge | 162 | failure to provide appropriate information to patient | 12 |
| substandard treatment | 311 | driving without tax, insurance, or licence caution | 161 | inadequate accessibility practice arrangements | 11 |
| no consent | 310 | driving without tax, insurance, or licence with no further action | 160 | poor response to audit or review | 10 |
| no consent to treatment | 309 | breach of the peace | 159 | failure to provide relevant information in references or reports | 9 |
| no consent to screening | 308 | breach of the peace conditional discharge | 158 | inefficient use of resources | 8 |
| exploiting patient financially or commercially | 307 | breach of the peace caution | 157 | unjustifiable comments in references or reports | 7 |
| pressure patient to go private | 306 | criminal damage | 156 | private dispute | 6 |
| UK regulatory determination | 305 | criminal damage conditional discharge | 155 | profession incidental not part of Good Medical Practice | 5 |
| overseas regulatory determination | 304 | criminal damage caution | 154 | issues cannot be identified | 4 |
| inadequate clinical skills | 303 | criminal damage with no further action | 153 | unregistered physician | 3 |
| inadequate history taking | 302 | resisting arrest | 152 | concern about the health system | 2 |
| inadequate assessment or history taking | 301 | resisting arrest caution | 151 | another healthcare professional | 1 |

For 47 investigations with no allegations, we assigned a seriousness rank of one, since 45 (96%) had no further action outcomes. To accommodate the potential natural absence of allegations elsewhere, we categorised seriousness ranks as: low [1, 150], medium (150, 300], and high (300, 450], plus a “no allegations” category. Ahead, we use “seriousness” as a shorthand for: the highest seriousness rank among all allegations considered in each decision.

## Results

In the 10 years ending with March 2023, out of 416,610 physicians registered in the UK, 8.8 % (36,685) had their first complaint triaged by the GMC. Fewer had first investigations (2.9 % or 12,237) and even fewer first hearings (0.5 % or 2,055).

### Severe outcomes related to serious concerns

Highly serious allegations were more common at further decision stages, making 55 % of first complaints with allegations, 63 % of first investigations, and 81 % of first hearings with proven allegations (table 1). Seriousness was highly related to promotion through stages. Complaints with the most serious allegations were investigated more often (fig. 1a; *n* = 262, Spearman *ρ* = 0.55, *p <* 0.0001), with only 3 % of complaints without allegations investigated (magenta). Investigation and hearing outcomes also related to seriousness (fig. 1b-e and 1f-i, *n* = 12, 237 and *n* = 2, 055, respectively, Kruskal-Wallis *p <* 0.0001). Highly serious allegations were more common for the most severe outcomes of referral to hearing and erasure (panels e and i).

**Figure 1:**
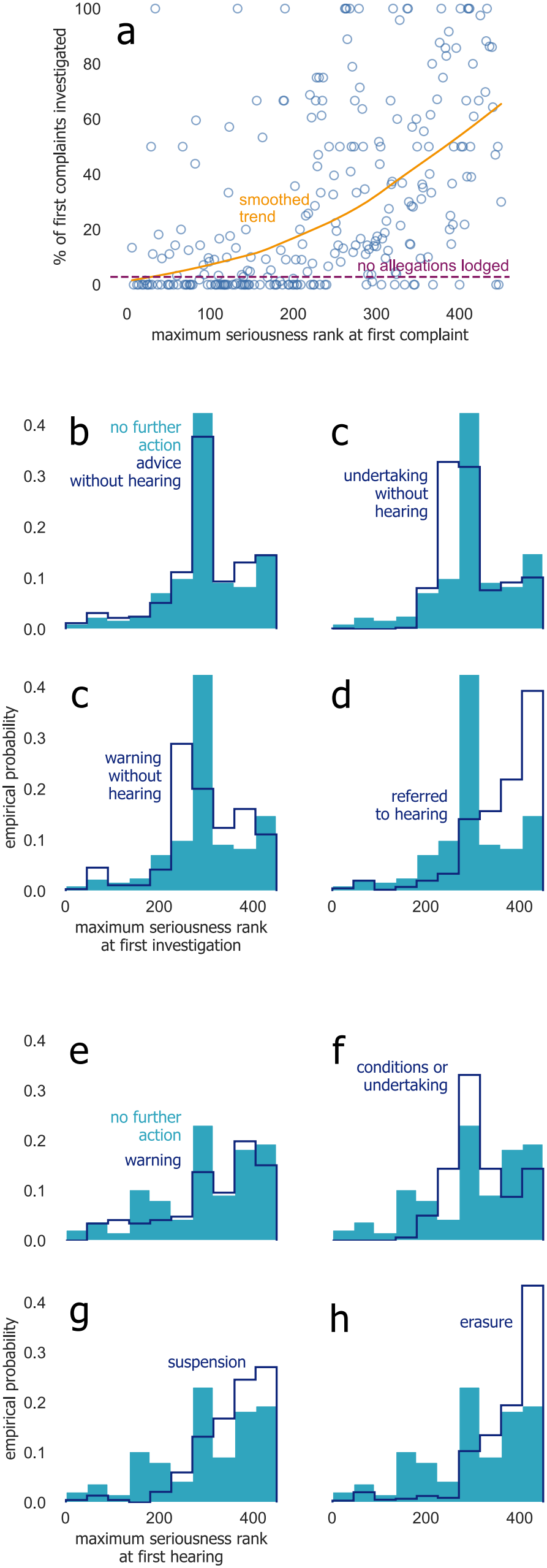
Seriousness related to decision outcomes. (a) Rate of investigation of first complaints against the rank of their most serious allegation since records of allegations at complaint stage started (blue), with a LOWESS smoothed trend (orange), and the investigation rate of complaints without allegations, thus seriousness information (magenta). (b-e) Distribution of the rank of the most serious allegation in all first investigations, by the outcome of the investigation (navy), contrasted to outcomes of no further action (teal). (f-i) Analogous to (b-e) but for first hearings.

Controlling for seriousness improved models of triage (AIC dropped from 30,369 in reduced model to 30,029), investigation (21,666 to 21,398), and hearing decisions (4,707 to 4,532), adding explanatory value to all (analysis of deviance, *p <* 0.0001).

Triage decisions tended to separate low-seriousness complaints from medium and high seriousness. Low seriousness strongly and consistently related to lowered chances of investigation (odds ratio (OR) 0.28; table 4), compared to medium seriousness. High seriousness related to increased chances (OR 1.46). Also, having no allegations, so no seriousness information, was unrelated to triage outcomes.

**Table 4:**
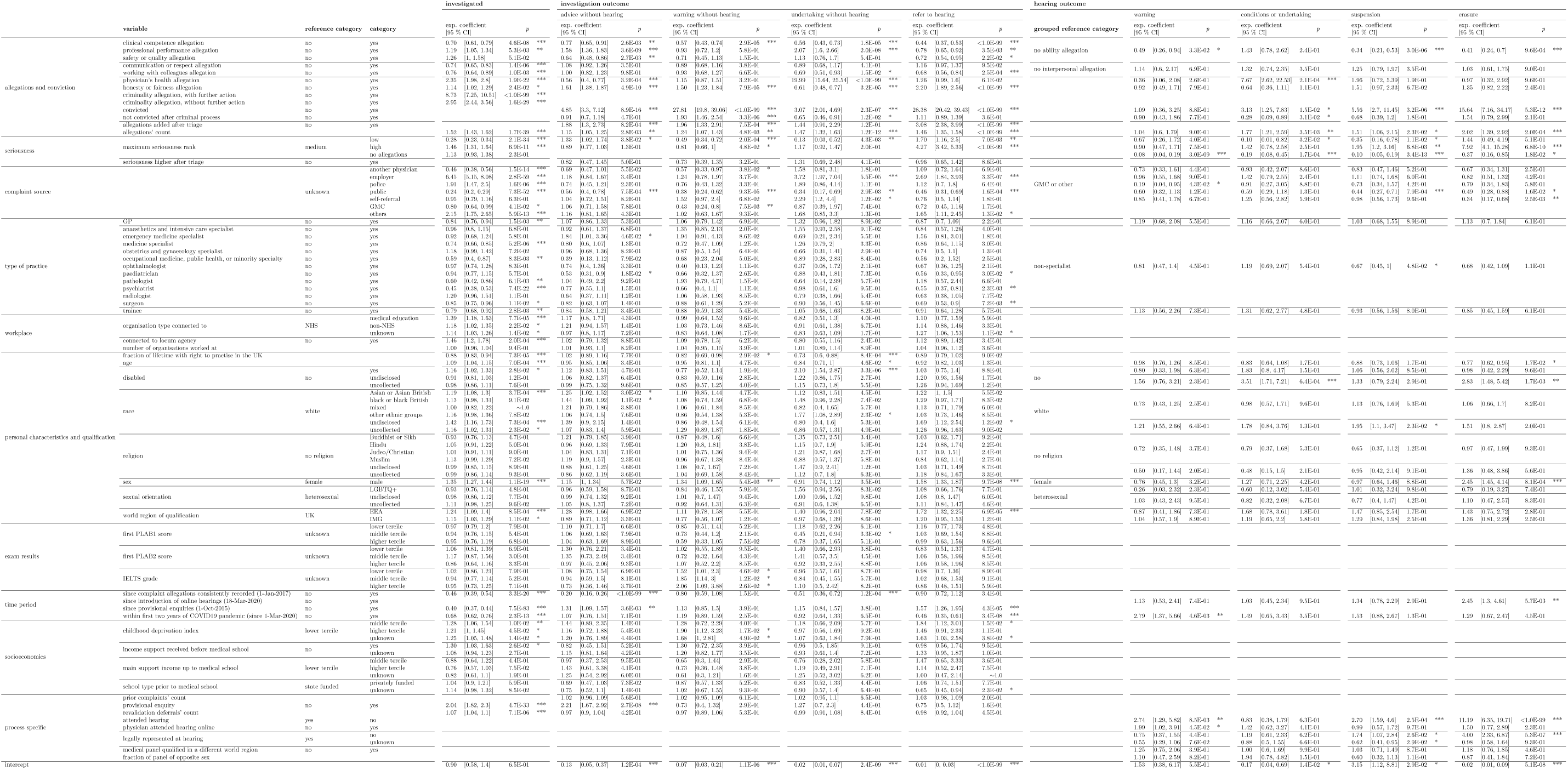
Results of modelling the three outcomes of: investigated (at triage), investigation outcome, and hearing outcome. Exponentiated coefficients [95 % confidence intervals], *p*-values in scientific notation, and significance level markers at 0.05 (*), 0.01 (**), and 0.001 (***). As in table 1, we further grouped categorical variables for the model of hearing outcomes.

|  |  |  |  | investigated |  | investigation outcome |  |  |  |  |  |  |  | hearing outcome |  |  |  |  |  |  |  |  |
|---|---|---|---|---|---|---|---|---|---|---|---|---|---|---|---|---|---|---|---|---|---|---|
|  |  |  |  |  |  | advice without hearing |  | warning without hearing |  | undertaking without hearing |  | refer to hearing |  |  | warning |  | conditions or undertaking |  | suspension |  | erasure |  |
|  | variable | reference category | category | exp. coefficient<br>[95 % CI] | p | exp. coefficient<br>[95 % CI] | p | exp. coefficient<br>[95 % CI] | p | exp. coefficient<br>[95 % CI] | p | exp. coefficient<br>[95 % CI] | p | grouped reference category | exp. coefficient<br>[95 % CI] | p | exp. coefficient<br>[95 % CI] | p | exp. coefficient<br>[95 % CI] | p | exp. coefficient<br>[95 % CI] | p |
| allegations and conviction | clinical competence allegation | no | yes | 0.70 [0.61, 0.79] | 4.6E-08 *** | 0.77 [0.65, 0.91] | 2.6E-03 ** | 0.57 [0.43, 0.74] | 2.9E-05 *** | 0.56 [0.43, 0.73] | 1.8E-05 *** | 0.44 [0.37, 0.53] | <1.0E-99 *** | no ability allegation | 0.49 [0.26, 0.94] | 3.3E-02 * | 1.43 [0.78, 2.62] | 2.4E-01 | 0.34 [0.21, 0.53] | 3.0E-06 *** | 0.41 [0.24, 0.7] | 9.6E-04 *** |
|  | professional performance allegation | no | yes | 1.19 [1.05, 1.34] | 5.3E-03 ** | 1.55 [1.36, 1.83] | 3.6E-09 *** | 0.93 [0.72, 1.2] | 5.8E-01 | 2.07 [1.6, 2.66] | 2.0E-08 *** | 0.78 [0.65, 0.92] | 3.5E-03 ** |  |  |  |  |  |  |  |  |  |
|  | safety or quality allegation | no | yes | 1.26 [1, 1.58] | 5.1E-02 | 0.64 [0.48, 0.86] | 2.7E-03 ** | 0.71 [0.45, 1.13] | 1.5E-01 | 1.13 [0.76, 1.7] | 5.4E-01 | 0.72 [0.54, 0.95] | 2.2E-02 * |  |  |  |  |  |  |  |  |  |
|  | communication or respect allegation | no | yes | 0.74 [0.65, 0.83] | 1.4E-06 *** | 1.08 [0.92, 1.26] | 3.5E-01 | 0.89 [0.68, 1.16] | 3.8E-01 | 0.89 [0.68, 1.17] | 4.1E-01 | 1.16 [0.97, 1.37] | 9.5E-02 | no interpersonal allegation | 1.14 [0.6, 2.17] | 6.9E-01 | 1.32 [0.74, 2.35] | 3.5E-01 | 1.25 [0.79, 1.97] | 3.5E-01 | 1.03 [0.61, 1.75] | 9.0E-01 |
|  | working with colleagues allegation | no | yes | 0.76 [0.64, 0.89] | 1.0E-03 ** | 1.00 [0.82, 1.23] | 9.8E-01 | 0.93 [0.68, 1.27] | 6.6E-01 | 0.69 [0.51, 0.93] | 1.5E-02 * | 0.68 [0.55, 0.84] | 2.5E-04 *** |  |  |  |  |  |  |  |  |  |
|  | physician's health allegation | no | yes | 2.35 [1.98, 2.8] | 1.9E-22 *** | 0.56 [0.4, 0.77] | 3.2E-04 *** | 1.15 [0.87, 1.51] | 3.2E-01 | 19.99 [15.64, 25.54] | <1.0E-99 *** | 1.26 [0.99, 1.6] | 6.1E-02 |  | 0.36 [0.06, 2.08] | 2.6E-01 | 7.67 [2.62, 22.53] | 2.1E-04 *** | 1.96 [0.72, 5.39] | 1.9E-01 | 0.97 [0.32, 2.92] | 9.6E-01 |
|  | honesty or fairness allegation | no | yes | 1.14 [1.02, 1.29] | 2.4E-02 * | 1.61 [1.38, 1.87] | 4.9E-10 *** | 1.50 [1.23, 1.83] | 7.9E-05 *** | 0.61 [0.48, 0.77] | 3.2E-05 *** | 2.20 [1.89, 2.56] | <1.0E-99 *** |  | 0.92 [0.49, 1.71] | 7.9E-01 | 0.64 [0.36, 1.11] | 1.1E-01 | 1.51 [0.97, 2.33] | 6.7E-02 | 1.35 [0.82, 2.22] | 2.4E-01 |
|  | criminality allegation, with further action | no | yes | 8.73 [7.25, 10.51] | <1.0E-99 *** |  |  |  |  |  |  |  |  |  |  |  |  |  |  |  |  |  |
|  | criminality allegation, without further action | no | yes | 2.95 [2.44, 3.56] | 1.6E-29 *** |  |  |  |  |  |  |  |  |  |  |  |  |  |  |  |  |  |
|  | convicted | no | yes |  |  | 4.85 [3.3, 7.12] | 8.9E-16 *** | 27.81 [19.8, 39.06] | <1.0E-99 *** | 3.07 [2.01, 4.69] | 2.3E-07 *** | 28.38 [20.42, 39.43] | <1.0E-99 *** |  | 1.09 [0.36, 3.25] | 8.8E-01 | 3.13 [1.25, 7.83] | 1.5E-02 * | 5.56 [2.7, 11.45] | 3.2E-06 *** | 15.64 [7.16, 34.17] | 5.3E-12 *** |
|  | not convicted after criminal process | no | yes |  |  | 0.91 [0.7, 1.18] | 4.7E-01 | 1.93 [1.46, 2.54] | 3.3E-06 *** | 0.65 [0.46, 0.91] | 1.2E-02 * | 1.11 [0.89, 1.39] | 3.6E-01 |  | 0.90 [0.43, 1.86] | 7.7E-01 | 0.28 [0.09, 0.89] | 3.1E-02 * | 0.68 [0.39, 1.2] | 1.8E-01 | 1.54 [0.79, 2.99] | 2.1E-01 |
|  | allegations added after triage | no | yes |  |  | 1.88 [1.3, 2.73] | 8.2E-04 *** | 1.96 [1.33, 2.91] | 7.5E-04 *** | 1.44 [0.92, 2.29] | 1.1E-01 | 3.08 [2.38, 3.99] | <1.0E-99 *** |  |  |  |  |  |  |  |  |  |
|  | allegations' count |  |  | 1.52 [1.43, 1.62] | 1.7E-39 *** | 1.15 [1.05, 1.25] | 2.8E-03 ** | 1.24 [1.07, 1.43] | 4.8E-03 ** | 1.47 [1.32, 1.63] | 1.2E-12 *** | 1.46 [1.35, 1.58] | <1.0E-99 *** |  | 1.04 [0.6, 1.79] | 9.0E-01 | 1.77 [1.21, 2.59] | 3.5E-03 ** | 1.51 [1.06, 2.15] | 2.3E-02 * | 2.02 [1.39, 2.92] | 2.0E-04 *** |
| seriousness | maximum seriousness rank | medium | low | 0.28 [0.23, 0.34] | 2.1E-34 *** | 1.33 [1.02, 1.74] | 3.8E-02 * | 0.49 [0.34, 0.72] | 2.0E-04 *** | 0.13 [0.03, 0.52] | 4.3E-03 ** | 1.70 [1.16, 2.5] | 7.0E-03 ** |  | 0.67 [0.26, 1.72] | 4.0E-01 | 0.10 [0.01, 0.82] | 3.2E-02 * | 0.35 [0.16, 0.78] | 1.1E-02 * | 1.44 [0.49, 4.19] | 5.1E-01 |
|  |  |  | high | 1.46 [1.31, 1.64] | 6.9E-11 *** | 0.89 [0.77, 1.03] | 1.3E-01 | 0.81 [0.66, 1] | 4.8E-02 * | 1.17 [0.92, 1.47] | 2.0E-01 | 4.27 [3.42, 5.33] | <1.0E-99 *** |  | 0.90 [0.47, 1.71] | 7.5E-01 | 1.42 [0.78, 2.58] | 2.5E-01 | 1.95 [1.2, 3.16] | 6.8E-03 ** | 7.92 [4.1, 15.28] | 6.8E-10 *** |
|  |  |  | no allegations | 1.13 [0.93, 1.38] | 2.3E-01 |  |  |  |  |  |  |  |  |  | 0.08 [0.04, 0.19] | 3.0E-09 *** | 0.19 [0.08, 0.45] | 1.7E-04 *** | 0.10 [0.05, 0.19] | 3.4E-13 *** | 0.37 [0.16, 0.85] | 1.8E-02 * |
|  | seriousness higher after triage | no | yes |  |  | 0.82 [0.47, 1.45] | 5.0E-01 | 0.73 [0.39, 1.35] | 3.2E-01 | 1.31 [0.69, 2.48] | 4.1E-01 | 0.96 [0.65, 1.42] | 8.6E-01 |  |  |  |  |  |  |  |  |  |
| complaint source |  | unknown | another physician | 0.46 [0.38, 0.56] | 1.5E-14 *** | 0.69 [0.47, 1.01] | 5.5E-02 | 0.57 [0.33, 0.97] | 3.8E-02 * | 1.58 [0.81, 3.1] | 1.8E-01 | 1.09 [0.72, 1.64] | 6.9E-01 | GMC or other | 0.73 [0.34, 1.61] | 4.4E-01 | 0.93 [0.42, 2.07] | 8.6E-01 | 0.83 [0.47, 1.46] | 5.2E-01 | 0.67 [0.34, 1.31] | 2.5E-01 |
|  |  |  | employer | 6.45 [5.15, 8.08] | 2.8E-59 *** | 1.18 [0.84, 1.67] | 3.4E-01 | 1.24 [0.78, 1.97] | 3.7E-01 | 3.72 [1.97, 7.04] | 5.5E-05 *** | 2.69 [1.84, 3.93] | 3.3E-07 *** |  | 0.96 [0.55, 1.68] | 9.0E-01 | 1.42 [0.79, 2.55] | 2.4E-01 | 1.11 [0.74, 1.68] | 6.0E-01 | 0.82 [0.51, 1.32] | 4.2E-01 |
|  |  |  | police | 1.91 [1.47, 2.5] | 1.6E-06 *** | 0.74 [0.45, 1.21] | 2.3E-01 | 0.76 [0.43, 1.32] | 3.3E-01 | 1.89 [0.86, 4.14] | 1.1E-01 | 1.12 [0.7, 1.8] | 6.4E-01 |  | 0.19 [0.04, 0.95] | 4.3E-02 * | 0.91 [0.27, 3.05] | 8.8E-01 | 0.73 [0.34, 1.57] | 4.2E-01 | 0.79 [0.34, 1.83] | 5.8E-01 |
|  |  |  | public | 0.24 [0.2, 0.29] | 7.3E-52 *** | 0.56 [0.4, 0.78] | 7.5E-04 *** | 0.38 [0.24, 0.62] | 9.3E-05 *** | 0.34 [0.17, 0.69] | 2.9E-03 ** | 0.46 [0.31, 0.69] | 1.6E-04 *** |  | 0.60 [0.32, 1.13] | 1.2E-01 | 0.59 [0.29, 1.18] | 1.3E-01 | 0.44 [0.27, 0.71] | 7.9E-04 *** | 0.49 [0.28, 0.88] | 1.6E-02 * |
|  |  |  | self-referral | 0.95 [0.79, 1.16] | 6.3E-01 | 1.04 [0.72, 1.51] | 8.2E-01 | 1.52 [0.97, 2.4] | 6.8E-02 | 2.29 [1.2, 4.4] | 1.2E-02 * | 0.76 [0.5, 1.14] | 1.8E-01 |  | 0.85 [0.41, 1.78] | 6.7E-01 | 1.25 [0.56, 2.82] | 5.9E-01 | 0.98 [0.56, 1.73] | 9.6E-01 | 0.34 [0.17, 0.68] | 2.5E-03 ** |
|  |  |  | GMC | 0.80 [0.64, 0.99] | 4.1E-02 * | 1.06 [0.71, 1.58] | 7.8E-01 | 0.43 [0.24, 0.8] | 7.5E-03 ** | 0.87 [0.39, 1.97] | 7.4E-01 | 0.72 [0.45, 1.16] | 1.7E-01 |  |  |  |  |  |  |  |  |  |
|  |  |  | others | 2.15 [1.75, 2.65] | 5.9E-13 *** | 1.16 [0.81, 1.65] | 4.3E-01 | 1.02 [0.63, 1.67] | 9.3E-01 | 1.68 [0.85, 3.3] | 1.3E-01 | 1.65 [1.11, 2.45] | 1.3E-02 * |  |  |  |  |  |  |  |  |  |
| type of practice | GP | no | yes | 0.84 [0.76, 0.94] | 1.5E-03 ** | 1.07 [0.86, 1.33] | 5.3E-01 | 1.06 [0.79, 1.42] | 6.9E-01 | 1.32 [0.96, 1.82] | 8.9E-02 | 0.87 [0.7, 1.09] | 2.2E-01 |  | 1.19 [0.68, 2.08] | 5.5E-01 | 1.16 [0.66, 2.07] | 6.0E-01 | 1.03 [0.68, 1.55] | 8.9E-01 | 1.13 [0.7, 1.84] | 6.1E-01 |
|  | anaesthetics and intensive care specialist | no | yes | 0.96 [0.8, 1.15] | 6.8E-01 | 0.92 [0.61, 1.37] | 6.8E-01 | 1.35 [0.85, 2.13] | 2.0E-01 | 1.55 [0.93, 2.58] | 9.1E-02 | 0.84 [0.57, 1.26] | 4.0E-01 | non-specialist | 0.81 [0.47, 1.4] | 4.5E-01 | 1.19 [0.69, 2.07] | 5.4E-01 | 0.67 [0.45, 1] | 4.8E-02 * | 0.68 [0.42, 1.09] | 1.1E-01 |
|  | emergency medicine specialist | no | yes | 0.92 [0.68, 1.24] | 5.8E-01 | 1.84 [1.01, 3.36] | 4.6E-02 * | 1.94 [0.91, 4.13] | 8.6E-02 | 0.69 [0.21, 2.34] | 5.5E-01 | 1.56 [0.81, 3.01] | 1.8E-01 |  |  |  |  |  |  |  |  |  |
|  | medicine specialist | no | yes | 0.74 [0.66, 0.85] | 5.2E-06 *** | 0.80 [0.6, 1.07] | 1.3E-01 | 0.72 [0.47, 1.09] | 1.2E-01 | 1.26 [0.79, 2] | 3.3E-01 | 0.86 [0.64, 1.15] | 3.0E-01 |  |  |  |  |  |  |  |  |  |
|  | obstetrics and gynaecology specialist | no | yes | 1.18 [0.99, 1.42] | 7.2E-02 | 0.96 [0.68, 1.36] | 8.2E-01 | 0.87 [0.5, 1.54] | 6.4E-01 | 0.66 [0.31, 1.41] | 2.9E-01 | 0.74 [0.5, 1.1] | 1.3E-01 |  |  |  |  |  |  |  |  |  |
|  | occupational medicine, public health, or minority specialty | no | yes | 0.59 [0.4, 0.87] | 8.3E-03 ** | 0.39 [0.13, 1.12] | 7.9E-02 | 0.68 [0.23, 2.04] | 5.0E-01 | 0.89 [0.28, 2.83] | 8.4E-01 | 0.56 [0.2, 1.52] | 2.5E-01 |  |  |  |  |  |  |  |  |  |
|  | ophthalmologist | no | yes | 0.97 [0.74, 1.28] | 8.3E-01 | 0.74 [0.4, 1.36] | 3.3E-01 | 0.40 [0.13, 1.23] | 1.1E-01 | 0.37 [0.08, 1.72] | 2.1E-01 | 0.67 [0.36, 1.25] | 2.1E-01 |  |  |  |  |  |  |  |  |  |
|  | paediatrician | no | yes | 0.94 [0.77, 1.15] | 5.7E-01 | 0.53 [0.31, 0.9] | 1.8E-02 * | 0.66 [0.32, 1.37] | 2.6E-01 | 0.88 [0.43, 1.81] | 7.3E-01 | 0.56 [0.33, 0.95] | 3.0E-02 * |  |  |  |  |  |  |  |  |  |
|  | pathologist | no | yes | 0.60 [0.42, 0.86] | 6.1E-03 ** | 1.04 [0.49, 2.2] | 9.2E-01 | 1.93 [0.79, 4.71] | 1.5E-01 | 0.64 [0.14, 2.99] | 5.7E-01 | 1.18 [0.57, 2.44] | 6.6E-01 |  |  |  |  |  |  |  |  |  |
|  | psychiatrist | no | yes | 0.45 [0.38, 0.53] | 7.4E-22 *** | 0.77 [0.55, 1.1] | 1.5E-01 | 0.66 [0.4, 1.1] | 1.1E-01 | 0.98 [0.61, 1.6] | 9.5E-01 | 0.55 [0.37, 0.81] | 2.3E-03 ** |  |  |  |  |  |  |  |  |  |
|  | radiologist | no | yes | 1.20 [0.96, 1.51] | 1.1E-01 | 0.64 [0.37, 1.11] | 1.2E-01 | 1.06 [0.58, 1.93] | 8.5E-01 | 0.79 [0.38, 1.66] | 5.4E-01 | 0.63 [0.38, 1.05] | 7.7E-02 |  |  |  |  |  |  |  |  |  |
|  | surgeon | no | yes | 0.85 [0.75, 0.96] | 1.1E-02 * | 0.82 [0.63, 1.07] | 1.4E-01 | 0.88 [0.61, 1.29] | 5.2E-01 | 0.90 [0.56, 1.45] | 6.6E-01 | 0.69 [0.53, 0.9] | 7.2E-03 ** |  |  |  |  |  |  |  |  |  |
|  | trainee | no | yes | 0.79 [0.68, 0.92] | 2.8E-03 ** | 0.84 [0.58, 1.21] | 3.4E-01 | 0.88 [0.59, 1.33] | 5.4E-01 | 1.05 [0.68, 1.63] | 8.2E-01 | 0.91 [0.64, 1.28] | 5.7E-01 |  | 1.13 [0.56, 2.26] | 7.3E-01 | 1.31 [0.62, 2.77] | 4.8E-01 | 0.93 [0.56, 1.56] | 8.0E-01 | 0.85 [0.45, 1.59] | 6.1E-01 |
| workplace | organisation type connected to | NHS | medical education | 1.39 [1.18, 1.63] | 7.7E-05 *** | 1.17 [0.8, 1.71] | 4.3E-01 | 0.99 [0.64, 1.52] | 9.6E-01 | 0.82 [0.51, 1.3] | 4.0E-01 | 1.10 [0.77, 1.59] | 5.9E-01 |  |  |  |  |  |  |  |  |  |
|  |  |  | non-NHS | 1.18 [1.02, 1.35] | 2.2E-02 * | 1.21 [0.94, 1.57] | 1.4E-01 | 1.03 [0.73, 1.46] | 8.6E-01 | 0.91 [0.61, 1.38] | 6.7E-01 | 1.14 [0.88, 1.46] | 3.3E-01 |  |  |  |  |  |  |  |  |  |
|  |  |  | unknown | 1.14 [1.03, 1.26] | 1.4E-02 * | 0.97 [0.8, 1.17] | 7.2E-01 | 0.83 [0.64, 1.08] | 1.7E-01 | 0.83 [0.63, 1.09] | 1.7E-01 | 1.27 [1.06, 1.53] | 1.1E-02 * |  |  |  |  |  |  |  |  |  |
|  | connected to locum agency | no | yes | 1.46 [1.2, 1.78] | 2.0E-04 *** | 1.02 [0.79, 1.32] | 8.8E-01 | 1.09 [0.78, 1.5] | 6.2E-01 | 0.80 [0.55, 1.16] | 2.4E-01 | 1.12 [0.89, 1.42] | 3.4E-01 |  |  |  |  |  |  |  |  |  |
|  | number of organisations worked at |  |  | 1.00 [0.96, 1.04] | 9.4E-01 | 1.01 [0.93, 1.1] | 8.2E-01 | 1.04 [0.95, 1.15] | 3.9E-01 | 1.01 [0.89, 1.14] | 8.9E-01 | 1.04 [0.96, 1.12] | 3.6E-01 |  |  |  |  |  |  |  |  |  |
| personal characteristics and qualification | fraction of lifetime with right to practise in the UK |  |  | 0.88 [0.83, 0.94] | 7.3E-05 *** | 1.02 [0.89, 1.16] | 7.7E-01 | 0.82 [0.69, 0.98] | 2.9E-02 * | 0.73 [0.6, 0.88] | 8.4E-04 *** | 0.89 [0.79, 1.02] | 9.0E-02 |  |  |  |  |  |  |  |  |  |
|  | age |  |  | 1.09 [1.04, 1.15] | 7.0E-04 *** | 0.95 [0.85, 1.06] | 3.4E-01 | 0.95 [0.81, 1.1] | 4.7E-01 | 0.84 [0.71, 1] | 4.6E-02 * | 0.92 [0.82, 1.03] | 1.3E-01 |  | 0.98 [0.76, 1.26] | 8.5E-01 | 0.83 [0.64, 1.08] | 1.7E-01 | 0.88 [0.73, 1.06] | 1.7E-01 | 0.77 [0.62, 0.95] | 1.7E-02 * |
|  | disabled | no | yes | 1.16 [1.02, 1.33] | 2.8E-02 * | 1.12 [0.83, 1.51] | 4.7E-01 | 0.77 [0.52, 1.14] | 1.9E-01 | 2.10 [1.54, 2.87] | 3.3E-06 *** | 1.03 [0.75, 1.4] | 8.8E-01 | no | 0.80 [0.33, 1.98] | 6.3E-01 | 1.83 [0.8, 4.17] | 1.5E-01 | 1.06 [0.56, 2.02] | 8.5E-01 | 0.98 [0.42, 2.29] | 9.6E-01 |
|  |  |  | undisclosed | 0.91 [0.81, 1.03] | 1.2E-01 | 1.06 [0.82, 1.37] | 6.4E-01 | 0.83 [0.59, 1.16] | 2.8E-01 | 1.22 [0.86, 1.75] | 2.7E-01 | 1.20 [0.93, 1.56] | 1.7E-01 |  | 1.56 [0.76, 3.21] | 2.3E-01 | 3.51 [1.71, 7.21] | 6.4E-04 *** | 1.33 [0.79, 2.24] | 2.9E-01 | 2.83 [1.48, 5.42] | 1.7E-03 ** |
|  |  |  | uncollected | 0.98 [0.86, 1.11] | 7.6E-01 | 0.99 [0.75, 1.32] | 9.6E-01 | 0.85 [0.57, 1.25] | 4.0E-01 | 1.15 [0.73, 1.8] | 5.5E-01 | 1.26 [0.94, 1.69] | 1.2E-01 |  |  |  |  |  |  |  |  |  |
|  | race | white | Asian or Asian British | 1.19 [1.08, 1.3] | 3.7E-04 *** | 1.25 [1.02, 1.52] | 3.0E-02 * | 1.10 [0.85, 1.44] | 4.7E-01 | 1.12 [0.83, 1.51] | 4.5E-01 | 1.22 [1, 1.5] | 5.5E-02 | white | 0.73 [0.43, 1.25] | 2.5E-01 | 0.98 [0.57, 1.71] | 9.6E-01 | 1.13 [0.76, 1.69] | 5.3E-01 | 1.06 [0.66, 1.7] | 8.2E-01 |
|  |  |  | black or black British | 1.13 [0.98, 1.31] | 9.1E-02 | 1.44 [1.09, 1.92] | 1.1E-02 * | 1.08 [0.74, 1.59] | 6.8E-01 | 1.48 [0.96, 2.28] | 7.4E-02 | 1.29 [0.97, 1.71] | 8.3E-02 |  |  |  |  |  |  |  |  |  |
|  |  |  | mixed | 1.00 [0.82, 1.22] | ~1.0 | 1.21 [0.79, 1.86] | 3.8E-01 | 1.06 [0.61, 1.84] | 8.5E-01 | 0.82 [0.4, 1.65] | 5.7E-01 | 1.13 [0.71, 1.79] | 6.0E-01 |  |  |  |  |  |  |  |  |  |
|  |  |  | other ethnic groups | 1.16 [0.98, 1.36] | 7.8E-02 | 1.06 [0.74, 1.5] | 7.6E-01 | 0.86 [0.54, 1.38] | 5.3E-01 | 1.77 [1.08, 2.89] | 2.3E-02 * | 1.03 [0.73, 1.46] | 8.5E-01 |  |  |  |  |  |  |  |  |  |
|  |  |  | undisclosed | 1.42 [1.16, 1.73] | 7.3E-04 *** | 1.39 [0.9, 2.15] | 1.4E-01 | 0.86 [0.48, 1.54] | 6.1E-01 | 0.80 [0.4, 1.6] | 5.3E-01 | 1.69 [1.12, 2.54] | 1.2E-02 * |  | 1.21 [0.55, 2.66] | 6.4E-01 | 1.78 [0.84, 3.76] | 1.3E-01 | 1.95 [1.1, 3.47] | 2.3E-02 * | 1.51 [0.8, 2.87] | 2.0E-01 |
|  |  |  | uncollected | 1.16 [1.02, 1.31] | 2.3E-02 * | 1.07 [0.83, 1.4] | 5.9E-01 | 1.29 [0.89, 1.87] | 1.8E-01 | 0.86 [0.57, 1.31] | 4.9E-01 | 1.26 [0.96, 1.63] | 9.0E-02 |  |  |  |  |  |  |  |  |  |
|  | religion | no religion | Buddhist or Sikh | 0.93 [0.76, 1.13] | 4.7E-01 | 1.21 [0.79, 1.85] | 3.9E-01 | 0.87 [0.48, 1.6] | 6.6E-01 | 1.35 [0.73, 2.51] | 3.4E-01 | 1.03 [0.62, 1.71] | 9.2E-01 | no religion | 0.72 [0.35, 1.48] | 3.7E-01 | 0.79 [0.37, 1.68] | 5.3E-01 | 0.65 [0.37, 1.12] | 1.2E-01 | 0.97 [0.47, 1.99] | 9.3E-01 |
|  |  |  | Hindu | 1.05 [0.91, 1.22] | 5.0E-01 | 0.96 [0.69, 1.33] | 7.9E-01 | 1.20 [0.8, 1.81] | 3.8E-01 | 1.15 [0.7, 1.9] | 5.9E-01 | 1.24 [0.88, 1.74] | 2.2E-01 |  |  |  |  |  |  |  |  |  |
|  |  |  | Judeo/Christian | 1.01 [0.91, 1.11] | 9.0E-01 | 1.04 [0.83, 1.31] | 7.1E-01 | 1.01 [0.75, 1.36] | 9.4E-01 | 1.21 [0.87, 1.68] | 2.7E-01 | 1.17 [0.9, 1.51] | 2.4E-01 |  |  |  |  |  |  |  |  |  |
|  |  |  | Muslim | 1.13 [0.99, 1.29] | 7.2E-02 | 1.19 [0.9, 1.57] | 2.3E-01 | 0.96 [0.67, 1.38] | 8.4E-01 | 0.88 [0.57, 1.35] | 5.8E-01 | 0.84 [0.62, 1.14] | 2.7E-01 |  |  |  |  |  |  |  |  |  |
|  |  |  | undisclosed | 0.99 [0.85, 1.15] | 8.9E-01 | 0.88 [0.61, 1.25] | 4.6E-01 | 1.08 [0.7, 1.67] | 7.2E-01 | 1.47 [0.9, 2.41] | 1.2E-01 | 1.03 [0.71, 1.49] | 8.7E-01 |  | 0.50 [0.17, 1.44] | 2.0E-01 | 0.48 [0.15, 1.5] | 2.1E-01 | 0.95 [0.42, 2.14] | 9.1E-01 | 1.36 [0.48, 3.86] | 5.6E-01 |
|  |  |  | uncollected | 0.99 [0.86, 1.14] | 9.3E-01 | 0.86 [0.62, 1.19] | 3.6E-01 | 1.04 [0.69, 1.58] | 8.4E-01 | 1.12 [0.7, 1.8] | 6.3E-01 | 1.18 [0.84, 1.67] | 3.3E-01 |  |  |  |  |  |  |  |  |  |
|  | sex | female | male | 1.35 [1.27, 1.44] | 1.1E-19 *** | 1.15 [1, 1.34] | 5.7E-02 | 1.34 [1.09, 1.65] | 5.4E-03 ** | 0.91 [0.74, 1.12] | 3.5E-01 | 1.58 [1.33, 1.87] | 9.7E-08 *** | female | 0.76 [0.45, 1.3] | 3.2E-01 | 1.27 [0.71, 2.25] | 4.2E-01 | 0.97 [0.64, 1.46] | 8.8E-01 | 2.45 [1.45, 4.14] | 8.1E-04 *** |
|  | sexual orientation | heterosexual | LGBTQ+ | 0.93 [0.76, 1.14] | 4.8E-01 | 0.96 [0.59, 1.58] | 8.7E-01 | 0.84 [0.46, 1.55] | 5.9E-01 | 1.56 [0.94, 2.56] | 8.3E-02 | 1.08 [0.66, 1.76] | 7.7E-01 | heterosexual | 0.26 [0.03, 2.32] | 2.3E-01 | 0.60 [0.12, 3.02] | 5.4E-01 | 1.01 [0.32, 3.24] | 9.8E-01 | 0.79 [0.19, 3.27] | 7.4E-01 |
|  |  |  | undisclosed | 0.98 [0.86, 1.12] | 7.7E-01 | 0.99 [0.74, 1.32] | 9.2E-01 | 1.01 [0.7, 1.47] | 9.4E-01 | 1.00 [0.66, 1.52] | 9.8E-01 | 1.08 [0.8, 1.47] | 6.0E-01 |  | 1.03 [0.43, 2.43] | 9.5E-01 | 0.82 [0.32, 2.08] | 6.7E-01 | 0.77 [0.4, 1.47] | 4.2E-01 | 1.10 [0.47, 2.57] | 8.3E-01 |
|  |  |  | uncollected | 1.11 [0.98, 1.25] | 9.6E-02 | 1.05 [0.8, 1.37] | 7.2E-01 | 0.92 [0.64, 1.31] | 6.3E-01 | 0.91 [0.6, 1.38] | 6.5E-01 | 1.11 [0.84, 1.47] | 4.6E-01 |  |  |  |  |  |  |  |  |  |
|  | world region of qualification | UK | EEA | 1.24 [1.09, 1.4] | 8.5E-04 *** | 1.28 [0.98, 1.66] | 6.9E-02 | 1.11 [0.78, 1.58] | 5.5E-01 | 1.40 [0.96, 2.04] | 7.8E-02 | 1.72 [1.32, 2.25] | 6.9E-05 *** |  | 0.87 [0.41, 1.86] | 7.3E-01 | 1.68 [0.78, 3.61] | 1.8E-01 | 1.47 [0.85, 2.54] | 1.7E-01 | 1.43 [0.75, 2.72] | 2.8E-01 |
|  |  |  | IMG | 1.15 [1.03, 1.29] | 1.1E-02 * | 0.89 [0.71, 1.12] | 3.3E-01 | 0.77 [0.56, 1.07] | 1.2E-01 | 0.97 [0.68, 1.39] | 8.6E-01 | 1.20 [0.95, 1.53] | 1.2E-01 |  | 1.04 [0.57, 1.9] | 8.9E-01 | 1.19 [0.65, 2.2] | 5.8E-01 | 1.29 [0.84, 1.98] | 2.5E-01 | 1.36 [0.81, 2.29] | 2.5E-01 |
| exam results | first PLAB1 score | unknown | lower tercile | 0.97 [0.79, 1.2] | 7.9E-01 | 1.10 [0.71, 1.7] | 6.6E-01 | 0.85 [0.51, 1.41] | 5.2E-01 | 1.18 [0.62, 2.26] | 6.1E-01 | 1.16 [0.77, 1.75] | 4.8E-01 |  |  |  |  |  |  |  |  |  |
|  |  |  | middle tercile | 0.94 [0.76, 1.15] | 5.4E-01 | 1.06 [0.69, 1.63] | 7.9E-01 | 0.73 [0.44, 1.2] | 2.1E-01 | 0.45 [0.21, 0.94] | 3.3E-02 * | 1.03 [0.69, 1.54] | 8.8E-01 |  |  |  |  |  |  |  |  |  |
|  |  |  | higher tercile | 0.95 [0.76, 1.19] | 6.8E-01 | 1.04 [0.63, 1.69] | 8.9E-01 | 0.59 [0.33, 1.05] | 7.5E-02 | 0.78 [0.37, 1.65] | 5.1E-01 | 0.99 [0.63, 1.56] | 9.6E-01 |  |  |  |  |  |  |  |  |  |
|  | first PLAB2 score | unknown | lower tercile | 1.06 [0.81, 1.39] | 6.9E-01 | 1.30 [0.76, 2.21] | 3.4E-01 | 1.02 [0.55, 1.89] | 9.5E-01 | 1.40 [0.66, 2.93] | 3.8E-01 | 0.83 [0.51, 1.37] | 4.7E-01 |  |  |  |  |  |  |  |  |  |
|  |  |  | middle tercile | 1.17 [0.87, 1.56] | 3.0E-01 | 1.35 [0.73, 2.49] | 3.4E-01 | 0.72 [0.32, 1.64] | 4.3E-01 | 1.41 [0.57, 3.5] | 4.5E-01 | 1.06 [0.58, 1.96] | 8.5E-01 |  |  |  |  |  |  |  |  |  |
|  |  |  | higher tercile | 0.86 [0.64, 1.16] | 3.3E-01 | 0.97 [0.45, 2.06] | 9.3E-01 | 1.07 [0.52, 2.2] | 8.5E-01 | 0.92 [0.33, 2.55] | 8.8E-01 | 1.06 [0.58, 1.96] | 8.5E-01 |  |  |  |  |  |  |  |  |  |
|  | IELTS grade | unknown | lower tercile | 1.02 [0.86, 1.21] | 7.9E-01 | 1.08 [0.75, 1.54] | 6.9E-01 | 1.52 [1.01, 2.3] | 4.6E-02 * | 0.96 [0.57, 1.61] | 8.7E-01 | 0.98 [0.7, 1.36] | 8.9E-01 |  |  |  |  |  |  |  |  |  |
|  |  |  | middle tercile | 0.94 [0.77, 1.14] | 5.2E-01 | 0.94 [0.59, 1.5] | 8.1E-01 | 1.85 [1.14, 3] | 1.2E-02 * | 0.84 [0.45, 1.55] | 5.7E-01 | 1.02 [0.68, 1.53] | 9.1E-01 |  |  |  |  |  |  |  |  |  |
|  |  |  | higher tercile | 0.95 [0.73, 1.25] | 7.1E-01 | 0.73 [0.36, 1.46] | 3.7E-01 | 2.06 [1.09, 3.88] | 2.6E-02 * | 1.10 [0.5, 2.42] | 8.2E-01 | 0.86 [0.48, 1.51] | 5.9E-01 |  |  |  |  |  |  |  |  |  |
| time period | since complaint allegations consistently recorded (1-Jan-2017) | no | yes | 0.46 [0.39, 0.54] | 3.3E-20 *** | 0.20 [0.16, 0.26] | <1.0E-99 *** | 0.80 [0.59, 1.08] | 1.5E-01 | 0.51 [0.36, 0.72] | 1.2E-04 *** | 0.90 [0.72, 1.12] | 3.4E-01 |  |  |  |  |  |  |  |  |  |
|  | since introduction of online hearings (18-Mar-2020) | no | yes |  |  |  |  |  |  |  |  |  |  |  | 1.13 [0.53, 2.41] | 7.4E-01 | 1.03 [0.45, 2.34] | 9.5E-01 | 1.34 [0.78, 2.29] | 2.9E-01 | 2.45 [1.3, 4.61] | 5.7E-03 ** |
|  | since provisional enquiries (1-Oct-2015) | no | yes | 0.40 [0.37, 0.44] | 7.5E-83 *** | 1.31 [1.09, 1.57] | 3.6E-03 ** | 1.13 [0.85, 1.5] | 3.9E-01 | 1.15 [0.84, 1.57] | 3.8E-01 | 1.57 [1.26, 1.95] | 4.3E-05 *** |  |  |  |  |  |  |  |  |  |
|  | within first two years of COVID19 pandemic (since 1-Mar-2020) | no | yes | 0.68 [0.62, 0.76] | 2.3E-13 *** | 1.07 [0.76, 1.51] | 7.1E-01 | 1.19 [0.89, 1.59] | 2.5E-01 | 0.92 [0.64, 1.33] | 6.5E-01 | 0.46 [0.35, 0.61] | 3.4E-08 *** |  | 2.79 [1.37, 5.66] | 4.6E-03 ** | 1.49 [0.65, 3.43] | 3.5E-01 | 1.53 [0.88, 2.67] | 1.3E-01 | 1.29 [0.67, 2.47] | 4.5E-01 |
| socioeconomics | childhood deprivation index | lower tercile | middle tercile | 1.28 [1.06, 1.54] | 1.0E-02 ** | 1.44 [0.89, 2.35] | 1.4E-01 | 1.28 [0.72, 2.29] | 4.0E-01 | 1.18 [0.66, 2.09] | 5.7E-01 | 1.84 [1.12, 3.01] | 1.5E-02 * |  |  |  |  |  |  |  |  |  |
|  |  |  | higher tercile | 1.21 [1, 1.45] | 4.5E-02 * | 1.16 [0.72, 1.88] | 5.4E-01 | 1.90 [1.12, 3.23] | 1.7E-02 * | 0.97 [0.56, 1.69] | 9.2E-01 | 1.46 [0.91, 2.33] | 1.1E-01 |  |  |  |  |  |  |  |  |  |
|  |  |  | unknown | 1.25 [1.05, 1.48] | 1.4E-02 * | 1.20 [0.76, 1.89] | 4.4E-01 | 1.68 [1, 2.81] | 4.9E-02 * | 1.07 [0.63, 1.84] | 7.9E-01 | 1.63 [1.03, 2.58] | 3.8E-02 * |  |  |  |  |  |  |  |  |  |
|  | income support received before medical school | no | yes | 1.30 [1.03, 1.63] | 2.6E-02 * | 0.82 [0.45, 1.51] | 5.2E-01 | 1.30 [0.72, 2.35] | 3.9E-01 | 0.96 [0.5, 1.85] | 9.1E-01 | 0.98 [0.56, 1.74] | 9.5E-01 |  |  |  |  |  |  |  |  |  |
|  |  |  | unknown | 1.08 [0.94, 1.23] | 2.7E-01 | 1.15 [0.81, 1.64] | 4.2E-01 | 1.20 [0.82, 1.77] | 3.5E-01 | 0.93 [0.61, 1.4] | 7.2E-01 | 1.33 [0.95, 1.87] | 1.0E-01 |  |  |  |  |  |  |  |  |  |
|  | main support income up to medical school | lower tercile | middle tercile | 0.88 [0.64, 1.22] | 4.4E-01 | 0.97 [0.37, 2.53] | 9.5E-01 | 0.65 [0.3, 1.44] | 2.9E-01 | 0.76 [0.28, 2.02] | 5.8E-01 | 1.47 [0.65, 3.33] | 3.6E-01 |  |  |  |  |  |  |  |  |  |
|  |  |  | higher tercile | 0.76 [0.57, 1.03] | 7.5E-02 | 1.43 [0.61, 3.38] | 4.1E-01 | 0.73 [0.36, 1.48] | 3.8E-01 | 1.19 [0.49, 2.91] | 7.1E-01 | 1.14 [0.52, 2.47] | 7.5E-01 |  |  |  |  |  |  |  |  |  |
|  |  |  | unknown | 0.82 [0.61, 1.1] | 1.9E-01 | 1.25 [0.54, 2.92] | 6.0E-01 | 0.61 [0.3, 1.21] | 1.6E-01 | 1.25 [0.52, 3.02] | 6.2E-01 | 1.00 [0.47, 2.14] | ~1.0 |  |  |  |  |  |  |  |  |  |
|  | school type prior to medical school | state funded | privately funded | 1.04 [0.9, 1.21] | 5.9E-01 | 0.69 [0.47, 1.03] | 7.3E-02 | 0.87 [0.57, 1.33] | 5.2E-01 | 0.83 [0.52, 1.33] | 4.4E-01 | 1.06 [0.74, 1.51] | 7.7E-01 |  |  |  |  |  |  |  |  |  |
|  |  |  | unknown | 1.14 [0.98, 1.32] | 8.5E-02 | 0.75 [0.52, 1.1] | 1.4E-01 | 1.02 [0.67, 1.55] | 9.3E-01 | 0.90 [0.57, 1.4] | 6.4E-01 | 0.65 [0.45, 0.94] | 2.3E-02 * |  |  |  |  |  |  |  |  |  |
| process specific | prior complaints' count |  |  |  |  | 1.02 [0.96, 1.09] | 5.6E-01 | 1.02 [0.95, 1.1] | 6.3E-01 | 1.02 [0.95, 1.09] | 6.5E-01 | 1.03 [0.98, 1.09] | 2.0E-01 |  |  |  |  |  |  |  |  |  |
|  | provisional enquiry | no | yes | 2.04 [1.82, 2.3] | 4.7E-33 *** | 2.21 [1.67, 2.92] | 2.7E-08 *** | 0.73 [0.4, 1.32] | 2.9E-01 | 1.27 [0.7, 2.3] | 4.3E-01 | 0.75 [0.5, 1.12] | 1.6E-01 |  |  |  |  |  |  |  |  |  |
|  | revalidation deferrals' count |  |  | 1.07 [1.04, 1.1] | 7.1E-06 *** | 0.97 [0.9, 1.04] | 4.2E-01 | 0.97 [0.89, 1.06] | 5.3E-01 | 0.99 [0.91, 1.08] | 8.4E-01 | 0.98 [0.92, 1.04] | 4.5E-01 |  |  |  |  |  |  |  |  |  |
|  | attended hearing | yes | no |  |  |  |  |  |  |  |  |  |  |  | 2.74 [1.29, 5.82] | 8.5E-03 ** | 0.83 [0.38, 1.79] | 6.3E-01 | 2.70 [1.59, 4.6] | 2.5E-04 *** | 11.19 [6.35, 19.71] | <1.0E-99 *** |
|  | physician attended hearing online | no | yes |  |  |  |  |  |  |  |  |  |  |  | 1.99 [1.02, 3.91] | 4.5E-02 * | 1.42 [0.62, 3.27] | 4.1E-01 | 0.99 [0.57, 1.72] | 9.7E-01 | 1.50 [0.77, 2.89] | 2.3E-01 |
|  | legally represented at hearing | yes | no |  |  |  |  |  |  |  |  |  |  |  | 0.75 [0.37, 1.55] | 4.4E-01 | 1.19 [0.61, 2.33] | 6.2E-01 | 1.74 [1.07, 2.84] | 2.6E-02 * | 4.00 [2.33, 6.87] | 5.3E-07 *** |
|  |  |  | unknown |  |  |  |  |  |  |  |  |  |  |  | 0.55 [0.29, 1.06] | 7.6E-02 | 0.88 [0.5, 1.55] | 6.6E-01 | 0.62 [0.41, 0.95] | 2.9E-02 * | 0.98 [0.58, 1.64] | 9.3E-01 |
|  | medical panel qualified in a different world region | no | yes |  |  |  |  |  |  |  |  |  |  |  | 1.25 [0.75, 2.06] | 3.9E-01 | 1.00 [0.6, 1.69] | 9.9E-01 | 1.03 [0.71, 1.49] | 8.7E-01 | 1.18 [0.76, 1.85] | 4.6E-01 |
|  | fraction of panel of opposite sex |  |  |  |  |  |  |  |  |  |  |  |  |  | 1.10 [0.47, 2.59] | 8.2E-01 | 1.94 [0.78, 4.82] | 1.5E-01 | 0.60 [0.32, 1.13] | 1.1E-01 | 0.87 [0.41, 1.84] | 7.2E-01 |
| intercept |  |  |  | 0.90 [0.58, 1.4] | 6.5E-01 | 0.13 [0.05, 0.37] | 1.2E-04 *** | 0.07 [0.03, 0.21] | 1.1E-06 *** | 0.02 [0.01, 0.07] | 2.4E-09 *** | 0.01 [0, 0.03] | <1.0E-99 *** |  | 1.53 [0.38, 6.17] | 5.5E-01 | 0.17 [0.04, 0.69] | 1.4E-02 * | 3.15 [1.12, 8.81] | 2.9E-02 * | 0.02 [0.01, 0.09] | 5.1E-08 *** |

High seriousness related to investigations ending in hearing referral (OR 4.27; table 4). Curiously, low seriousness also appeared related to hearing referral, barely within our chosen significance level, which we interpret as a large-sample artefact (OR 1.70, *n* = 12, 237, *p* = 0.007). Low seriousness related to lower chances of warning or undertaking (ORs 0.49 and 0.13). Seriousness appeared unrelated to outcomes of advice without a hearing. Lastly, increased seriousness after triage, via allegations added during investigation, was unrelated to investigation outcomes.

Highly serious proven allegations at hearing were strongly related to erasure (OR 7.92; table 4), compared to medium seriousness. Low seriousness strongly related to closure with no further action, instead of suspension, or conditions or undertaking (ORs 0.35 or 0.10). Having no allegations proven, so no seriousness information, strongly related to lower chances of erasure, suspension, conditions or under-taking, or warning (ORs 0.37, 0.10, 0.19, or 0.08).

### Seriousness mediated the importance of allegations

Allegation types coded concern themes and related to seriousness (fig. 2a; *n* = 684, Kruskal-Wallis *p <* 0.0001). However, within types, allegation codes can differ widely in seriousness; take the criminal matters of murder and driving without insurance (top of fig. 2a). Hence, controlling for seriousness gave a better-adjusted association for allegation types.

**Figure 2:**
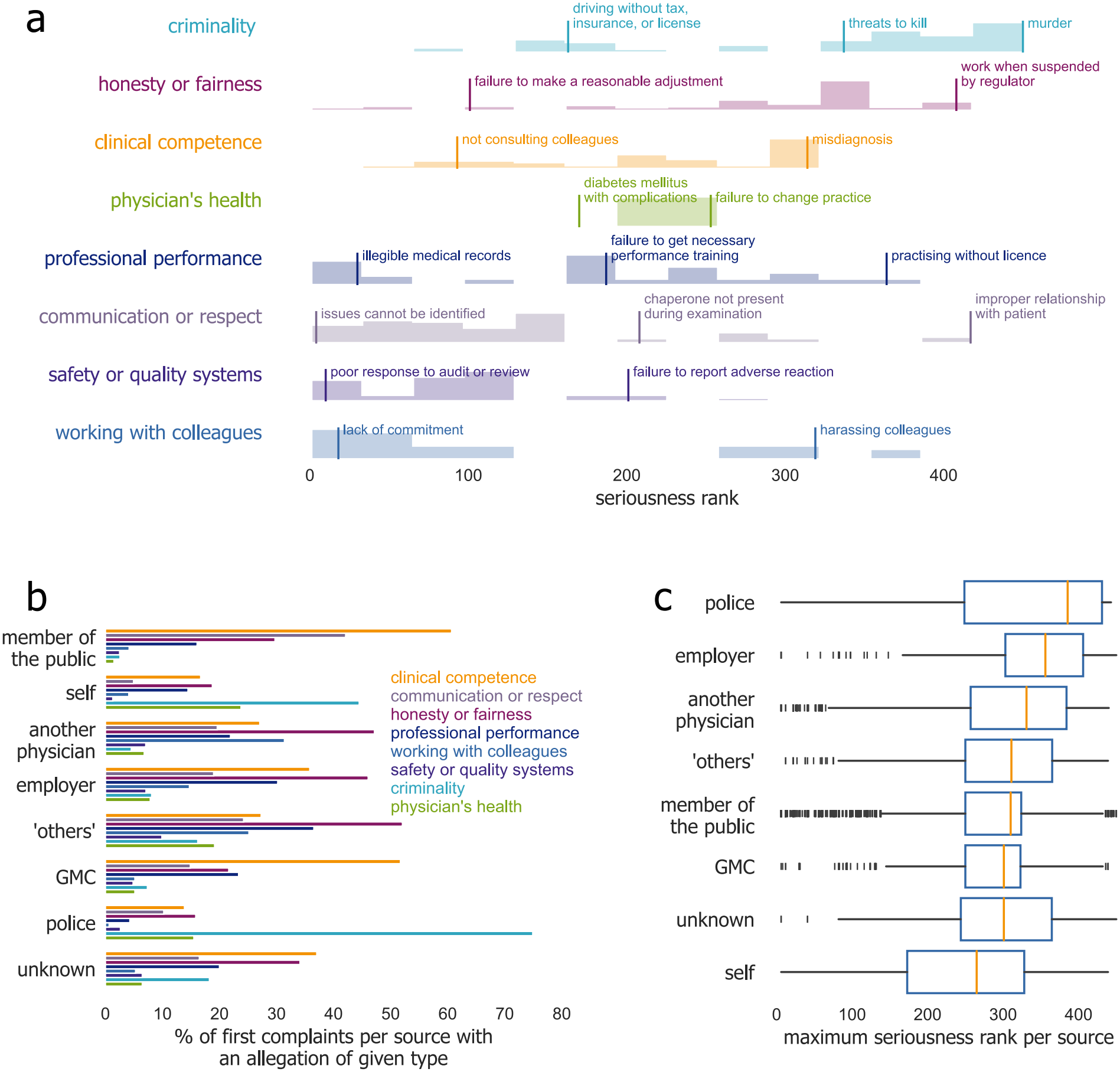
Association between allegation type, seriousness, and complaint source. (a) Distribution of the 684 allegation codes by allegation type (criminality consolidated), with example allegation codes labelled at their rank. (b) Distribution of allegation types per source among first complaints; groups ordered by the prevalence of complaint sources. (c) Distribution of seriousness of first complaints by complaint source (quartiles; whiskers span 1.5 times the interquartile range). Data for b-c since 1-Jan-2017, when allegation records at the complaint stage started.

The prevalence of allegation types differed by complaint source (fig. 2b; *n* = 12, 237, Fisher’s exact *p <* 0.0001 for every type), and seriousness was more similar between sources (panel c; though Kruskal-Wallis *p <* 0.0001, *n* = 12, 237). Hence, source ORs remained relatively unchanged after controlling for seriousness across all decisions, compared to reduced models.

#### Seriousness was critical to nuance criminality involvement

For sanctions, seriousness was crucial in appraising the appropriate importance of conviction; suggested by controlling for seriousness substantially shrinking the effect size of conviction for sanctions of erasure, suspension, or conditions or undertaking at hearing (OR from 28.61 in reduced model to 15.64, 10.01 to 5.56, or 4.74 to 3.13, respectively).

Seriousness enhanced clarifying that alleged criminality was separately important in prompting further scrutiny. Controlling for seriousness amplified the effect size of allegations of criminality, both with and without further action, for investigation at triage (OR 6.85 in reduced model to 8.73, table 4; and from 2.27 to 2.95, respectively). The same happened for conviction and outcomes of referral to hearing after investigation (from OR 21.79 in reduced model to 28.38).

#### Seriousness upstaged physician’s health allegations

Controlling for seriousness absorbed variability from allegations about the physician’s health, regarding complaints being investigated (OR decreased from 2.71 in reduced model to 2.35; table 4). Also, regarding outcomes of undertaking without hearing (OR decreased from 21.14 to 19.99), and outcomes of conditions or undertaking after hearing (from 15.32 to 7.67). This upstaging was partly because health allegations clustered around medium seriousness ranks, and were seldom the most serious (fig. 2a).

#### Seriousness explained away associations with some allegation types

Allegations of honesty or fairness were among the highest-effect factors consistently associated with a complaint being investigated in the reduced triage model (OR 1.44). The same was apparent regarding outcomes of erasure, suspension, or warning in the reduced model of hearing outcomes (ORs 5.17, 5.30, and 2.59, respectively). Allegations of communication, respect, or working with colleagues were also among the top factors in the latter reduced model, associated with suspension or warning outcomes (ORs of 2.45 and 2.25). All such effect sizes were substantially reduced and became non-significant after controlling for seriousness (table 4), suggesting that the importance of such allegations operated primarily through how serious concerns were.

### Alongside seriousness, conviction, allegation types, and source were the main factors related to outcomes

Complaint investigation at triage was primarily related to: allegations about criminality or the physician’s health; being a complaint by the employer, police, or others acting in a professional capacity (‘others’ category), relative to unknown source; becoming a “provisional enquiry;” more allegations; and high seriousness, relative to medium (table 4). Complaint closure with no further action mainly related to: originating from the public or another physician; low seriousness; originating since the implementation of either provisional enquiries or pre-triage allegation recording, highlighting the value of such information-enhancing mechanisms; and specialising in psychiatry, pathology, occupational medicine, public health, and/or a minority specialty.

Referral to hearing, the most severe investigation outcome, principally related to: criminal conviction; high seriousness, relative to medium; adding allegations during investigation; being a complaint by the employer, relative to unknown source; honesty or fairness allegations; and qualifying in the EEA, relative to the UK (table 4), which we hypothesise hinges on lower engagement with the process, as seen at hearings. ^18^ Conversely, lower chances of a referral to hearing mostly related to: clinical competence allegations; being a complaint from the public; and being a psychiatrist. Conviction strongly associated with receiving all other investigation outcomes instead of no further action. Undertaking without a hearing chiefly related to physician’s health or professional performance allegations, to being an employer complaint, and to disability, reflecting an undertaking’s aim to keep a physician safely in practice.

Receiving the most severe outcome of erasure after a hearing predominantly related to: conviction; not attending the hearing (congruent with ^18^); high seriousness, relative to medium; and lack of legal representation (table 4). Suspension was also primarily related to conviction and non-attendance. Lowering chances of erasure was mainly related to self-referral and having no proven allegations —hence no seriousness information. Lowering the chances of suspension was also mainly related to having no proven allegations, followed by proven clinical competence, professional performance, or safety or quality systems allegations, low seriousness, and being a public complaint. As with undertaking without hearing, conditions or undertaking after a hearing were chiefly related to physician’s health allegations, followed by an unknown disability status, and conviction. Low seriousness strongly related to lowering the chances of suspension, or conditions or undertaking. Interestingly, attending their hearing online was unrelated to the physician receiving a sanction, but related to increased chances of a warning. This suggests that the efficiencies a “virtual hearing” brings to all involved could be safely considered by other jurisdictions.

## Discussion

We asked whether regulatory decisions about physicians related to the seriousness of the concerns raised. Collaborating with decision experts, we created a new measure of seriousness. We showed that the factors chiefly related to decisions were consistent across decision stages: criminal conviction, seriousness, allegation types, and the complaint’s source. This agrees with precedents regarding allegation types and sources, ^10,18^ while meaningfully refining current understanding of akin decisions by bringing in conviction and seriousness.

The main limitation of this study was focusing on UK regulatory decisions related to medical practice. Regardless, we believe our findings can generalise to meaningfully inform wider research and policy in similar jurisdictions in other geographies and professions. The main strength of this study was its design. We examined regulatory decision stages individually and separately from complainants’ decisions, thereby avoiding confounders from complaint motivations. Our models fitted notably well and rendered large adjusted ORs, which is uncommon when modelling real-world data. We hypothesise this is attributable to extensive written policies, guidance, and legal frame-works standardising decision-making.

Another strength was the great number and variety of independent variables studied, contrasting with precedents’ limitation to few variables; ^1–4,8–13,16,19,21,34–39^ most commonly, in order, sex, area of practice, domestic or international qualification, practise length and type, allegations, age, and race, focusing on characteristics that are non-modifiable for risk mitigation. Our richer context helped us pinpoint which factors predominantly related to decisions and, just as crucially, which were not. This addressed many live questions —only a few discussed at length. Nonetheless, as in any similar design, there still could be important unidentified latent variables. Some are anecdotally reported to affect akin decisions, but remain hard to measure and therefore analyse quantitatively. For example: acceptance of accountability, contrition or insight, candour, and remediation. ^20,40^

Large sample sizes enabled us to study rare but important factors —like conviction or qualifying in certain specialties. Also, to strongly suggest that factors with non-significant adjusted ORs were unrelated to decision outcomes, *e*.*g*. sexual orientation or religion. However, large samples complicate the interpretation of hypothesis-testing results. Especially for factors with low but significant adjusted effect sizes. For example, a desirable statistical power of ~80 % for the association of being Asian and being investigated at triage (OR 1.19, or an expected probability of investigation increased by two percentage points, from the average marginal effect), required 70 % of the sample used by our triage model (*n* = 25, 679; *post-hoc* simulation drawing random samples without replacement). Large-sample requirements like these question the reliability of akin small-effect-size associations, despite statistical significance — and support interpreting such associations as artefacts of working with large samples.

Personal characteristics tended to be unrelated to regulatory decisions: inconsistent associations despite large samples, relatively small effect sizes, and/or artefacts of large samples. This questions assumptions that physicians from certain demographic groups practise less safely. Nonetheless, lingering associations could mask unidentified confounders that could guide productive interventions. ^18^ Especially regarding being male, which is often found to relate to severe disciplinary, legal, and regulatory outcomes worldwide, ^2,3,5,10,15–17,21,35,37,39,41–46^ without clear underlying reasons why. ^16,47^

### The difficulty of measuring seriousness

Seriousness is an abstract concept. It is uncountable and has no units of measurement, leading to many methods to quantify it; all subject to common methodological issues, *e*.*g*. whether to consider historical decision outcomes, multiple concerns underlying each outcome, or multiple people involved. ^33^ Addressing more issues better nuances a measure, but also complicates it, obstructing its sustainable practical use. ^28^ We kept our seriousness ranking simple to avoid most methodological issues while allowing meaningful analysis. Future applications may require more complexity.

Our ranking is an ordinal scale of allegation-code locations, implying equal distances between adjacent codes. One could weight such distances using data on concern prevalence and outcomes, as in criminal indices using police data. ^27–33^ However, such circular reasoning ^22^ would hinder asking whether serious concerns beget severe outcomes, as we did.

Variability around allegation code locations also matters. Additional factors ^22^ (some of which we controlled for) determine the seriousness of cases sharing a code, hence why jurisdictions appraise each case in its own context. ^20^ For example, murder allegations are highly serious, with little variability across individual cases. Conversely, misdiagnosis is of medium seriousness, but could vary widely —sometimes causing minor discomfort, others late-stage cancer detection.

We anticipated some association between our seriousness ranking and outcomes, since decision experts helped develop it. Nonetheless, we believe our ranking matches well current societal expectations of physicians, making it useful in its context. Supporting our ranking’s validity was, first, that having no allegations was unrelated to triage outcomes, and strongly related to no further action outcomes after hearing. Second, that only experts in triage and investigation decisions collaborated on its development, yet the ranking was also highly explanatory of hearing decisions — a form of cross-validation.

## Conclusion

Seriousness of concerns related to severe regulatory outcomes, alongside other main factors like criminal conviction, allegations of criminality, honesty, or fairness, and the complaint source being the physician’s employer, the police, or others acting in a professional capacity. Unlike precedents, our large-scale study found that personal characteristics had little to no association with outcomes. This underlines what physicians are pragmatically expected to avoid in their day-to-day, to keep patients safe and maintain trust in the medical profession.

## Data Availability

The data analysed includes personally identifiable information on sensitive processes and characteristics, entrusted to the GMC. To protect these, data is only available upon request in aggregated, anonymised formats like those in the report.

## Contributors

The author conceptualised, designed and ran this study, coordinated the engineering of the datasets, conducted the literature review and statistical analysis, coordinated quality assurance exercises, wrote all drafts, and had final responsibility for the decision to submit for publication.

## Ethical approval

Not required.

## Acknowledgements

We want to thank Ken Leach, David Finan, Laura Cashin, and Sav Tyrimos for their input. Elizabeth Monks, Dyvia Mishra, Adrian McAndrew, Bushra Hussain, and Harry Barton, among others, for data engineering. Also, David Darton, Joanna Farrell, Colin Melville, Nathan Booth, Mason Tiffany, and James Gooding for useful comments on earlier manuscripts.

## References

[1] Clay SW, Conatser RR. Characteristics of physicians disciplined by the State Medical Board of Ohio. Journal of the American Osteopathic Association. 2003;103(2):81–8.

[2] Kohatsu ND, Gould D, Ross LK, Fox PJ. Characteristics associated with physician discipline: a case-control study. Archives of internal medicine. 2004 Mar;164:653–8.

[3] Khaliq AA, Dimassi H, Huang CY, Narine L, Smego RA. Disciplinary action against physicians: who is likely to get disciplined? The American journal of medicine. 2005 Jul;118:773–7.

[4] Cardarelli R, Licciardone JC. Factors Associated With High-Severity Disciplinary Action by a State Medical Board: A Texas Study of Medical License Revocation. The Journal of the American Osteopathic Association. 2006;106(3):153–6.

[5] Tamblyn R, Abrahamowicz M, Dauphinee D, Wenghofer E, Jacques A, Klass D, et al. Physician scores on a national clinical skills examination as predictors of complaints to medical regulatory authorities. JAMA. 2007 Sep;298(9):993–1001.

[6] Pukk-Härenstam K, Ask J, Brommels M, Thor J, Penaloza RV, Gaffney FA. Analysis of 23 364 patient-generated, physician-reviewed malpractice claims from a non-tort, blame-free, national patient insurance system: lessons learned from Sweden. Postgraduate medical journal. 2009 Feb;85:69–73.

[7] Wu CY, Lai HJ, Chen RC. Medical malpractice experience of Taiwan: 2005 versus 1991. Internal medicine journal. 2009 Apr;39:237–42.

[8] Alam A, Klemensberg J, Griesman J, Bell CM. The characteristics of physicians disciplined by professional colleges in Canada. Open medicine. 2011;5(4):e166–72.

[9] Elkin KJ, Spittal MJ, Elkin DJ, Studdert DM. Doctors disciplined for professional misconduct in Australia and New Zealand, 2000-2009. The Medical journal of Australia. 2011 May;194(9):452–6.

[10] Humphrey C, Hickman S, Gulliford MC. Place of medical qualification and outcomes of UK General Medical Council “fitness to practise” process: cohort study. BMJ. 2011 Apr;342:d1817.

[11] Alam A, Kurdyak P, Klemensberg J, Griesman J, Bell CM. The characteristics of psychiatrists disciplined by professional colleges in Canada. PloS ONE. 2012;7:e50558.

[12] Elkin K, Spittal MJ, Studdert DM. Risks of complaints and adverse disciplinary findings against international medical graduates in Victoria and Western Australia. The Medical journal of Australia. 2012 Oct;197(8):448–52.

[13] Birkeland S, Depont Christensen R, Damsbo N, Kragstrup J. Characteristics of complaints resulting in disciplinary actions against Danish GPs. Scandinavian journal of primary health care. 2013 Sep;31:153–7.

[14] Birkeland S, Christensen Rd, Damsbo N, Kragstrup J. Process-related factors associated with disciplinary board decisions. BMC health services research. 2013 Jan;13:9.

[15] Spittal MJ, Bismark MM, Studdert DM. The PRONE score: an algorithm for predicting doctors’ risks of formal patient complaints using routinely collected administrative data. BMJ Qual Saf. 2015;24(6):360–8.

[16] Unwin E, Woolf K, Wadlow C, Potts HWW, Dacre J. Sex differences in medico-legal action against doctors: a systematic review and meta-analysis. BMC medicine. 2015 Aug;13:172.

[17] Studdert DM, Bismark MM, Mello MM, Singh H, Spittal MJ. Prevalence and Characteristics of Physicians Prone to Malpractice Claims. The New England journal of medicine. 2016 Jan;374(4):354–62.

[18] Caballero JA, Brown SP. Engagement, not personal characteristics, was associated with the seriousness of regulatory adjudication decisions about physicians: a cross-sectional study. BMC medicine. 2019;17:1–10.

[19] Sampieri G, Xu J, Noel CW, Matelski J, Liu JJ, Bell CM, et al. Surgeons disciplined by regulatory bodies in Canada between 2000 and 2017. Canadian journal of surgery Journal canadien de chirurgie. 2022;65:E656–60.

[20] Bryce M, Reynolds E, Price T, Quick O, OBŕien T, Endacott R, et al. The concept of seriousness in fitness to practise cases. University of Plymouth; 2022.

[21] Wakeford R. Who gets struck off? BMJ. 2011 Dec;343:d7842.

[22] Bryce M, Archer J, Brennan N, Burns L, OBŕien T, Price T. Fitness to practise: Impairment and Serious Misconduct, A narrative synthesis review. University of Plymouth; 2018.

[23] Ridley S. Severity of illness scoring systems and performance appraisal. Anaesthesia. 1998 Dec;53:1185–94.

[24] Bouch DC, Thompson JP. Severity scoring systems in the critically ill. Continuing Education in Anaesthesia Critical Care & Pain. 2008 10;8(5):181–5.

[25] Sellin T, Wolfgang ME. The Measurement of Delinquency. Wiley; 1964.

[26] Wolfgang ME MFR, Tracy PE, Singer SI. The National Survey of Crime Severity. U.S. Department of Justice, Bureau of Justice Statistics; 1985.

[27] Wallace M, Turner J, Matarazzo A, Babyak C. Measuring crime in Canada: Introducing the Crime Severity Index and improvements to the Uniform Crime Reporting survey. Ottawa: Statistics Canada; 2009. No. 85-004-X.

[28] Sherman L, Neyroud PW, Neyroud E. The Cambridge Crime Harm Index: Measuring Total Harm from Crime Based on Sentencing Guidelines. Policing: A Journal of Policy and Practice. 2016 04;10(3):171–83.

[29] Bangs M. Research outputs: developing a Crime Severity Score for England and Wales using data on crimes recorded by the police. Newport: Office for National Statistics. 2016.

[30] Curtis-Ham S, Walton D. The New Zealand crime harm index: Quantifying harm using sentencing data. Policing: A Journal of Policy and Practice. 2018;12(4):455–67.

[31] House PD, Neyroud PW. Developing a crime harm index for Western Australia: The WACHI. Cambridge Journal of Evidence-Based Policing. 2018;2:70–94.

[32] Kärrholm F, Neyroud P, Smaaland J. Designing the Swedish Crime Harm Index: an evidence-based strategy. Cambridge Journal of Evidence-Based Policing. 2020;4:15–33.

[33] van Ruitenburg T, Ruiter S. The adoption of a crime harm index: A scoping literature review. Police Practice and Research. 2023;24(4):423–45.

[34] Dehlendorf CE, Wolfe SM. Physicians disciplined for sex-related offenses. JAMA. 1998 Jun;279:1883–8.

[35] Morrison J, Wickersham P. Physicians disciplined by a state medical board. JAMA. 1998 Jun;279(23):1889–93.

[36] Morrison J, Morrison T. Psychiatrists disciplined by a state medical board. The American journal of psychiatry. 2001 Mar;158(3):474–8.

[37] National Clinical Assessment Service. NCAS casework: the first eight years. National Health Service; 2009.

[38] National Clinical Assessment Service. Concerns about professional practice and associations with age, gender, place of qualification and ethnicity - 2009/10 data. National Health Service; 2010.

[39] Unwin E, Woolf K, Wadlow C, Dacre J. Disciplined doctors: Does the sex of a doctor matter? A cross-sectional study examining the association between a doctor’s sex and receiving sanctions against their medical registration. BMJ Open. 2014;4:e005405.

[40] Johnstone MJ, Kanitsaki O. Processes for disciplining nurses for unprofessional conduct of a serious nature: a critique. Journal of advanced nursing. 2005 May;50:363–71.

[41] Cooil B. Using Medical Malpractice Data to Predict the Frequency of Claims: A Study of Poisson Process Models with Random Effects. Journal of the American Statistical Association. 1991;86(414):285–95.

[42] Gibbons RD, Hedeker D, Charles SC, Frisch P. A random-effects probit model for predicting medical malpractice claims. Journal of the American Statistical Association. 1994;89(427):760–7.

[43] Weycker DA, Jensen GA. Medical malpractice among physicians: Who will be sued and who will pay? Health Care Management Science. 2000 Sep;3(4):269–77.

[44] Hickson GB, Federspiel CF, Pichert JW, Miller CS, Gauld-Jaeger J, Bost P. Patient complaints and malpractice risk. JAMA. 2002 Jun;287(22):2951–7.

[45] Rolph JE, Adams JL, McGuigan KA. Identifying Malpractice-Prone Physicians. Journal of Empirical Legal Studies. 2007;4(1):125–53.

[46] Taragin MI, Wilczek AP, Karns ME, Trout R, Carson JL. Physician demographics and the risk of medical malpractice. The American journal of medicine. 1992 Nov;93:537–42.

[47] Foong-Reichert AL, Fung A, Carter CA, Grindrod KA, Houle SK. Characteristics, predictors and reasons for regulatory body disciplinary action in health care: a scoping review. Journal of Medical Regulation. 2021;107(4):17–31.

